# A Decade of Progress in HIV, Malaria, and Tuberculosis Initiatives in Malawi

**DOI:** 10.1101/2024.10.08.24315077

**Authors:** Tara Danielle Mangal, Margherita Molaro, Dominic Nkhoma, Timothy Colbourn, Joseph H. Collins, Eva Janoušková, Matthew M. Graham, Ines Li Lin, Emmanuel Mnjowe, Tisungane E. Mwenyenkulu, Sakshi Mohan, Bingling She, Asif U. Tamuri, Pakwanja D. Twea, Peter Winskill, Andrew Phillips, Joseph Mfutso-Bengo, Timothy B. Hallett

## Abstract

**Objective:** Huge investments in HIV, TB, and malaria (HTM) control in Malawi have greatly reduced disease burden. However, the joint impact of these services across multiple health domains and the health system resources required to deliver them are not fully understood.

**Methods:** An integrated epidemiological and health system model was used to assess the impact of HTM programmes in Malawi from 2010 to 2019, incorporating interacting disease dynamics, intervention effects, and health system usage. Four scenarios were examined, comparing actual programme delivery with hypothetical scenarios excluding programmes individually and collectively.

**Findings:** From 2010-2019, HTM programmes were estimated to have prevented 1.08 million deaths and 74.89 million DALYs. An additional 15,600 deaths from other causes were also prevented. Life expectancy increased by 13.0 years for males and 16.9 years for females.

The HTM programmes accounted for 24.2% of all health system interactions, including 157.0 million screening/diagnostic tests and 23.2 million treatment appointments. Accounting for the anticipated health deterioration without HTM services, only 41.55 million additional healthcare worker hours were required (17.1% of total healthcare worker time) to achieve these gains. The HTM programme eliminated the need for 123 million primary care appointments, offset by a net increase in inpatient care demand (9.4 million bed-days) that would have been necessary in its absence.

**Conclusions:** HTM programmes have greatly increased life expectancy, providing direct and spillover effects on health. These investments have alleviated the burden on inpatient and emergency care, which requires more intensive healthcare provider involvement.

## 1 Introduction

Between 2010 and 2020, Malawi’s substantial investments in HIV/AIDS, tuberculosis (TB) and malaria (HTM) programmes have significantly reduced disease burden.[1] This decade of targeted interventions saw remarkable progress in public health through comprehensive testing, treatment, and preventive services.

Widespread access to antiretroviral therapy (ART) for HIV has improved life expectancy and health for individuals living with HIV in Malawi. The adoption of the Joint United Nations programme on HIV/AIDS (UNAIDS) 90-90-90 targets resulted in a 71% decrease in new HIV infections between 2010 and 2020, dropping to 17,000 annually, and a 65% reduction in AIDS deaths, falling to 13,000.[2] Prevention of mother-to-child transmission services reached 96.3% for maternal ART and 92.3% for infant prophylaxis by 2021.[3] TB control efforts have improved case detection rates from 42% in 2010 to 56% in 2020, while the TB treatment success rate for drug-sensitive strains has increased from 73% to 85%.[4] Insecticide-treated bednets and effective antimalarial drugs have substantially reduced malaria morbidity and mortality. In 2020, malaria incidence had decreased to 219 cases per 1,000 people, from 381 cases per 1,000 people in 2010. Similarly, the malaria mortality rate dropped to 38 deaths per 100,000 people, compared to 73 deaths per 100,000 people in 2010.[5]

While the direct health benefits of these programmes are clear, their broader health system implications and spillover effects are less explored.[6-8] For instance, controlling HIV infections can lead to reductions in risks associated with diarrhoeal disease [9], acute lower respiratory illness (ALRI) [10], childhood under-nutrition and stunting [11], non-AIDS cancers [12], cardiovascular and cerebrovascular disease [13], depression [14] and maternal anaemia [15] (see Figure 1). Similarly, malaria control efforts can reduce the risk of conditions like maternal anaemia, stillbirth, and preterm birth.[16] Additionally, TB control has been linked to a decrease in the incidence and severity of diabetes.[17] Therefore HTM programmes may have far-reaching health impacts beyond their primary targets, potentially influencing healthcare utilisation and overall system demand.

**Figure 1:**
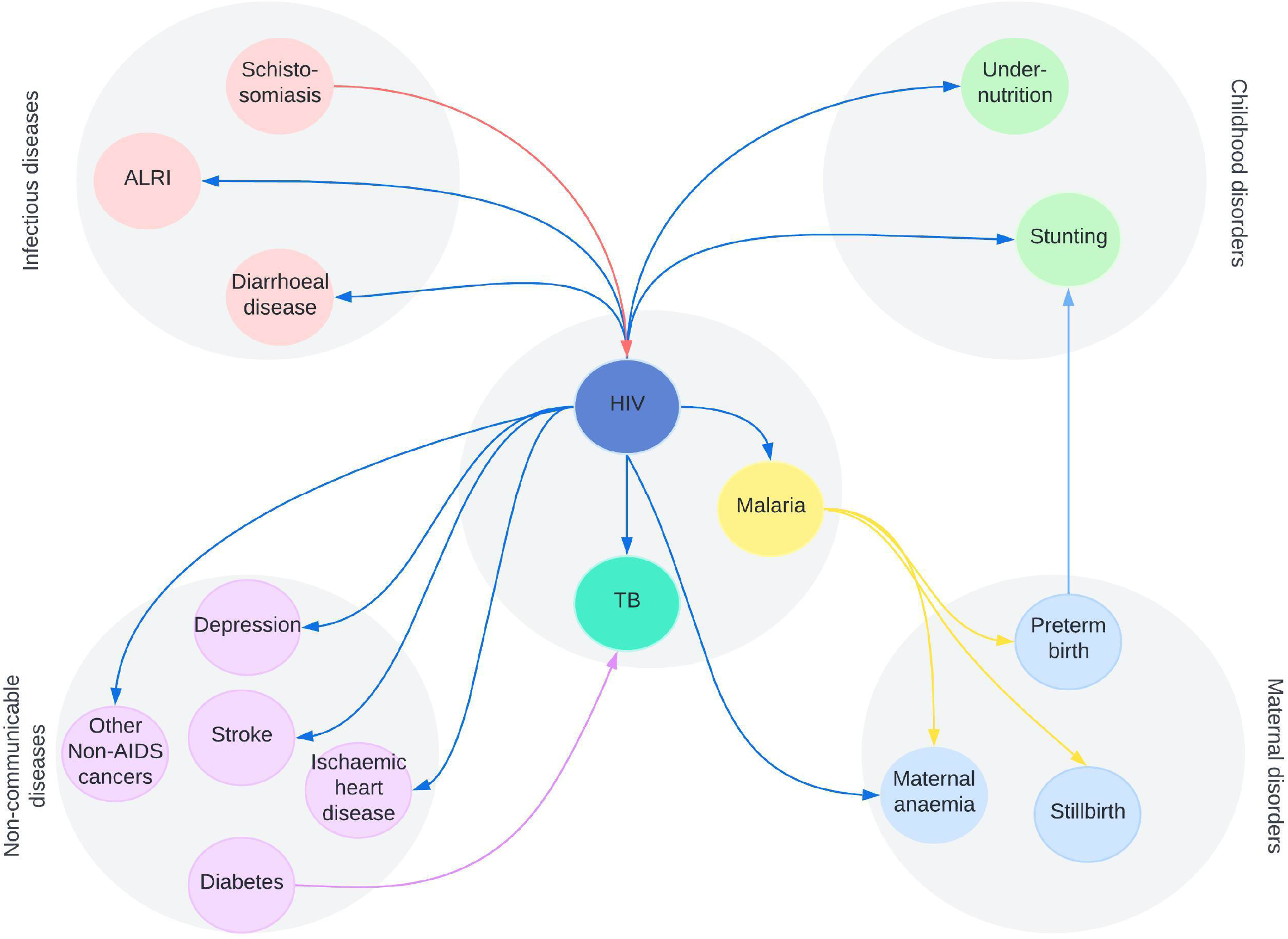
The biological interactions for HIV, TB and malaria captured in the Thanzi la Onse model. Arrows depict the direction of risk, e.g. HIV is a contributing cause for diarrhoeal disease and acute lower respiratory infections (ALRI) etc. Social and behavioural risk factors are not depicted here.

The evaluation of global health programmes traditionally relies on disease-specific models, focusing solely on their individual health impacts.[6, 18-20] This siloed approach, however, has limitations. Firstly, it cannot capture the broader benefits of these interventions on other health conditions. Secondly, the siloed approach doesn’t account for individuals with multiple infections who might be counted towards “deaths averted” in multiple programmes. This can lead to an inflated sense of programme effectiveness, as someone potentially saved from one disease might still succumb to another within the same time-frame.

To address these limitations, health systems modelling is gaining traction, considering the real-world complexity of healthcare systems and allowing for the evaluation of programmes with a wider perspective.[21] It can quantify the programme’s impact on the entire spectrum of health conditions and healthcare system utilisation. Additionally, it can assess the overall healthcare system needs associated with HTM programs, both in the context of programme implementation and inaction.

As Malawi begins extensive health system reforms, strengthening existing health system capabilities, addressing bottlenecks and integrating service delivery, it becomes evident that a comprehensive health system modelling framework is required that can provide a panoramic view of the healthcare landscape.[22, 23]

Here we use a whole health-system model to quantify the impact that HTM programmes have had in Malawi between 2010 and 2019 by i) estimating the combined programme effects in terms of direct health benefits; ii) estimating the spillover effects into other health conditions; iii) quantifying the demands on the health system required to achieve these health benefits and iv) simulating the hypothetical demands on the health system had the three programmes not operated.

## 2 Methods

### 2.1 TLO model

This analysis uses the Thanzi la Onse model, a dynamic whole health-system model simulating the lifetime health of a representative Malawian population cohort.[24] The model, detailed online with accessible source code, has been validated against reported data on disease burden, health system engagement, key consumables availability, and service delivery volume.[25-28] It includes three core features: a realistic representation of the Malawian health system, a simulated population facing lifetime health hazards, and a statistical model of health system engagement. Disease modules within the model track the onset, progression, and care outcomes, with certain diseases, such as HIV and malaria, contributing to others like TB and maternal anaemia.

Each simulated individual has attributes like sex, residence, education, wealth quintile, and lifestyle factors (smoking, alcohol use) based on DHS surveys, which can change over time.[27] Detailed contraception choices, pregnancy, labor, and delivery are modeled and calibrated to match available data.[29] When ill or pregnant, individuals may seek healthcare immediately or after a delay, depending on symptoms, sex, age, wealth, and residence. The model predicts the likelihood of seeking care using estimates from Ng’ambi et al. (2020).[30, 31]

#### 2.1.1 Health system capabilities

The health system’s capabilities depend on healthcare personnel distribution, availability of essential medicines and diagnostic tools, and hospital bed capacity.[26] Individuals can seek care at any district facility, influenced by their symptoms and characteristics, with care provided according to the Malawi Clinical Guidelines.[32] In the model, appointments can cover multiple services, such as TB screening and malaria treatment, with patient-facing time quantified for each cadre. The availability of key medicines determines if appointments can proceed; if not, patients may return later or default. Healthcare services are delivered via community programs, outpatient clinics, or district/referral hospitals, with “outpatient appointments” encompassing all these interactions.

#### 2.1.2 Disease modules

The TLO framework models health conditions responsible for 81% of deaths and 72% of disability-adjusted life-years (DALYs) in Malawi from 2015 to 2019.[33] The disease models interact with each other and with individual attributes to represent, for example, shared risk factors for multiple diseases and the influence of one disease on the risk or severity of another. Each disease model has been calibrated to specific data with guidance from programme managers and disease experts. These disease models, calibrated with input from experts, account for interactions between diseases, shared risk factors, and the impact of one disease on another. The framework covers neonatal and early childhood conditions, maternal conditions, communicable and non-communicable diseases, and injuries, with additional risks modelled to align with Global Burden of Disease estimates. These disease models, calibrated with input from experts, account for interactions between diseases, shared risk factors, and the impact of interventions across multiple health conditions. The framework covers neonatal and early childhood conditions, maternal conditions, communicable and non-communicable diseases, and injuries, with additional risks modelled to align with Global Burden of Disease estimates. The analysis focuses on three diseases, detailed further in Mangal *et al* and the Supplementary Information.[34]

##### HIV

HIV risk in adults is modelled based on sexual contact, with varying risk factors including sex, wealth, education, VMMC, PrEP use, or commercial sex work for females. Annual acquisition risk is calibrated using MPHIA and UNAIDS data (2010-2022). Mother-to-child transmission is modelled across pregnancy, labour, and breastfeeding, influenced by maternal ART and infant prophylaxis.

HIV testing can occur via provider-initiated testing during other clinic visits, self-initiated testing, antenatal clinics, or routine testing for PrEP users. Positive results prompt a referral for treatment, with viral suppression determined at treatment initiation. Individuals may stop or default on ART (due to stockouts), seek re-initiation later, or undergo retesting as needed. Negative test results prompt a referral to other appropriate services such as behaviour change counselling, VMMC or PrEP.

##### Tuberculosis

Active TB infections are determined based on untreated TB prevalence and individual risk factors such as BCG vaccination, obesity, smoking, HIV status, and Isoniazid preventive therapy (IPT) usage. Drug-sensitive and multidrug resistant (MDR) TB strains are modelled separately. TB relapse risk is higher for people living with HIV (PLHIV), and mortality is linked to smear status and treatment success, occurring 1-5 months after onset. Incidence and mortality estimates are aligned with WHO (2022) and Malawi NTP reports (2018, 2019).

Diagnostic tools include sputum smear microscopy, GeneXpert, chest X-ray, and clinical diagnosis, each with varying sensitivity and specificity. GeneXpert is prioritised for PLHIV and relapse cases, chest X-ray is required for children under 5, and clinical diagnosis is used if other tests are unavailable. Positive results prompt immediate treatment, with MDR-TB cases receiving specialised regimens. Annual treatment coverage is aligned with Malawi NTP reports (2017-2021). IPT is given routinely to reduce active TB risk, with a 6-month course for TB contacts and a 36-month regimen for PLHIV starting IPT with ART.

##### Malaria

The natural history of malaria is captured through an emulation model, based on an individual based simulation calibrated to parasite prevalence data.[35] This model captures age-specific, district-level incidences of asymptomatic, clinical, and severe malaria in the presence of interventions such as indoor residual spraying and long-lasting insecticide-treated bednets. It reflects the full dynamics of *P. falciparum* transmission and incorporates age and exposure-dependent immunity functions. By emulating this transmission model, we account for complex factors including malaria seasonality, the decay of maternal immunity in the first six months of life, and the acquired immunity that develops with parasite exposure, which varies with transmission intensity. Intervention coverage is informed by data from The Malaria Atlas Project.[36]

Symptom onset typically occurs 7 days after infection, with clinical cases resolving through treatment or self-cure. Severe cases require emergency care and are subject to mortality risks based on treatment status. The risks of clinical and severe malaria are heightened in individuals with unsuppressed HIV infections, the effects of which can be mitigated through antiretroviral therapy (ART) and Cotrimoxazole use.

Rapid diagnostic tests are used to confirm malaria diagnoses for all patients presenting with fever and are also offered through community outreach programs to both symptomatic and asymptomatic individuals. Treatment is tailored to clinical severity, effectively resolving symptoms and clearing parasitaemia within 7 days. Pregnant women attending antenatal care (ANC) services are provided intermittent preventive treatment of malaria (IPTp), which confers 6 weeks of protection against clinical disease.

##### Disease interactions

The TLO model incorporates a number of disease interactions relating to HIV, TB and malaria, detailed in Figure 1 and the Supplementary Information. Many conditions additionally share risk factors, for example excessive alcohol use can exacerbate the risk or severity of cardiometabolic disorders, TB and mental health (not shown in Figure). Additional indirect links exist, for example malaria increases the risk of preterm labour, leading to stunting in children, a known risk factor for diarrhoeal diseases. Integrated healthcare approaches are included in the model, such as HIV testing offered through TB clinics, or bednets and IPTp given through antenatal clinics, reflecting the standard Malawi Clinical Guidelines.[32]

### 2.2 Model simulations

The model is initialised in 2010 with a representative simulated population of 147,000 (1:100) and runs for 10 years. Individuals are assigned demographic characteristics (age, sex, parity) and attributes related to lifestyle (education, wealth, obesity, smoking), health (non-communicable and infectious diseases, mental health, reproductive or newborn health), and prior healthcare use (treatment status, vaccination history). All code is executed in Python programming language 3.8.[37] Five model runs are performed for each scenario, with medians, and 95% uncertainty intervals calculated for each output.

#### Definition of scenarios

We simulate the “Actual” scenario, depicting the services that were delivered throughout 2010–2019, along with four hypothetical counterfactual scenarios where HTM service packages are excluded individually or in combination for the duration of the simulation (Table 1). Under the hypothetical scenarios, when the HIV/TB/malaria services are excluded, only end-of-life or palliative care is provided for HIV, TB and malaria patients. A summary of the health status of the population and health system usage was produced every year and DALYs were calculated using the disability weights from Salomon *et al*.[38] To calculate life expectancy at birth, standard life table methods were employed using 5-year age groups up to age 90+ and adjusted to separately account for age-groups <1 year and 1-4 years.[39]

**Table 1:** Description of the services included in each scenario for the period 2010–2019. All non-HTM services are consistently available as required across the scenarios, reflecting the true service availability throughout this period. Average coverage values in eligible populations are given for 2019 but vary by district and year throughout the simulation. Cotrimoxazole is given to people with HIV to prevent opportunistic infections; here we consider only the effect on malaria incidence/severity. Abbreviations: PMTCT prevention of mother-to-child transmission; FSW female sex workers.

| Scenario | Description | Coverage in 2019 |
| --- | --- | --- |
| Actual | All services are included | Antiretroviral therapy: 85% in adults, 100% in children<br>PMTCT: 95%<br>Pre-exposure prophylaxis for FSW: 5%<br>Infant prophylaxis: 80%<br>Voluntary medical male circumcision: 30%<br>TB treatment: 75%<br>TB preventive therapy: 67%<br>BCG vaccination: 91%<br>Malaria treatment: 42%<br>Insecticide-treated bednets: 79%<br>Indoor residual spraying: 5%<br>Intermittent preventive therapy for pregnant women (2 doses): 76%<br>Cotrimoxazole: 89% |
| No HIV Services | As for actual but with no HIV services available | Antiretroviral therapy: 0%<br>PMTCT: 0%<br>Pre-exposure prophylaxis: 0%<br>Infant prophylaxis: 0%<br>Voluntary medical male circumcision: 0% |
| No TB Services | As for actual but with no TB services available | TB treatment: 0%<br>TB preventive therapy: 0%<br>Contact investigation for index cases: None<br>BCG vaccination: 0% |
| No Malaria Services | As for actual but with no malaria services available | Malaria treatment: 0%<br>Insecticide-treated bednets: 0%<br>Indoor residual spraying: 0%<br>Intermittent preventive therapy for pregnant women: 0%<br>Cotrimoxazole: 0% |
| No HTM Services | As for actual but with no HTM services available | Antiretroviral therapy: 0%<br>PMTCT: 0%<br>Pre-exposure prophylaxis: 0%<br>Infant prophylaxis: 0%<br>Voluntary medical male circumcision: 0%<br>TB treatment: 0%<br>TB preventive therapy: 0%<br>BCG vaccination: 0%<br>Malaria treatment: 0%<br>Insecticide-treated bednets: 0%<br>Indoor residual spraying: 0%<br>Intermittent preventive therapy for pregnant women: 0%<br>Cotrimoxazole: 0% |

#### Ethical approval

The Thanzi La Onse project received ethical approval from the College of Medicine Malawi Research Ethics Committee (COMREC, P.10/19/2820) for the use of publicly accessible and anonymised secondary data. No data were used requiring individual informed consent.

## 3 Results

### 3.1 The health impacts of each programme

#### Direct health benefits

Excluding each set of services in turn and comparing population health with the Actual scenario gives us an estimate of the health gains attributable to each programme. The provision of HIV-related healthcare services is estimated to have averted 31.95 million DALYs (95% UI 31.60–32.74 million) due to HIV/AIDS and 261,100 (95% UI 135,900– 417,100) DALYs due to TB during 2010– 2019 (see Supplementary Information). TB services have directly prevented 5.48 million (5.32 - 6.06 million) TBDALYs, with a further 1.04 million (95% UI 573,6000–1,55 million) DALYs averted due to HIV/AIDS. The provision of malaria services averted 36.84 million (95% UI 35.94– 37.25 million) DALYs due to malaria.

HTM programmes (when considered in combination) have prevented 579,300 (95% UI 570,900–586,100), 94,200 (90,400–100,900) and 416,100 (414,000–420,800) deaths due to HIV/AIDS, TB and malaria respectively and 74.89 million DALYs (74.18–75.16 million) HTM DALYs over a ten year period (Figure 2), increasing life expectancy at birth in 2019 for women by 16.9 years (from 50.4 to 66.1 years) and men by 13.0 years (from 48.2 to 61.7 years).

**Figure 2:**
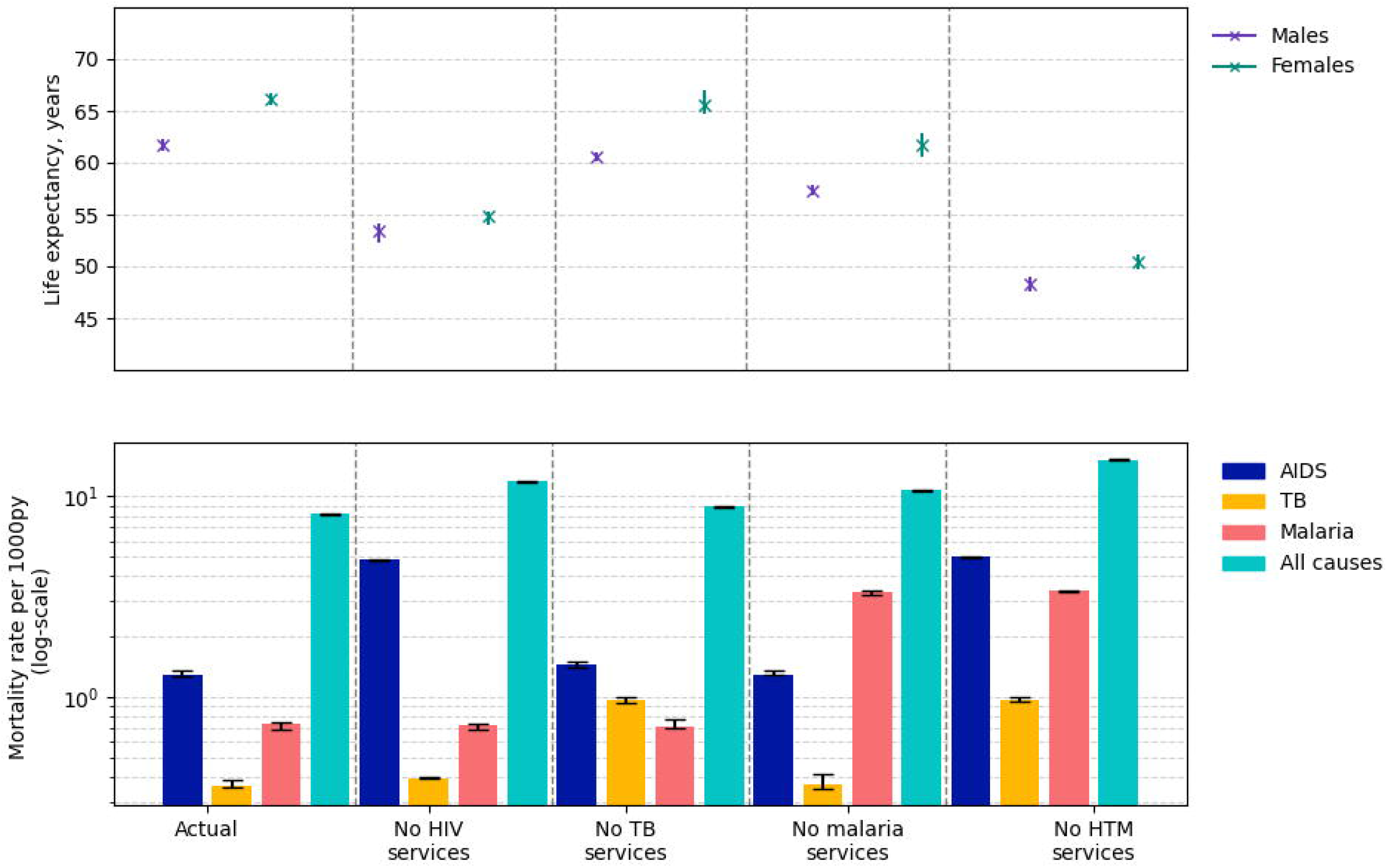
The estimated life expectancy in years from birth in 2019 for males and females by scenario (top figure) and the mortality rates per 1000 person-years due to HIV/AIDS, TB, malaria and all causes by scenario (lower figure). The error bars show the 95% uncertainty intervals across the 5 runs of each draw.

#### Spillover effects on other diseases

Our findings reveal substantial reductions in the incidence of childhood diarrhoea, ALRI, non-AIDS cancers and TB associated with HIV services, averting an estimated 1.99 million DALYs (95% UI 1.37–2.31 million DALYs associated with these conditions). Likewise, both TB and malaria services have had an impact on other health conditions, with reductions in DALY burden attributed to AIDS (1.04 million DALYs averted) and ALRI (140,700 DALYs averted) associated with TB services and improvements in diabetes (42,200 DALYs averted) and neonatal disorders (337,100 DALYs averted) attributed to malaria services. The assessment of overall health benefits stemming from HTM services is complicated by shifting demographics (see Supplementary Figures). One such effect is the reduction of mortality rates, leading to a rise in person-years at risk for developing other conditions such as COPD, diabetes, stroke, and kidney disease. Jointly, HTM services have averted 15,600 deaths (95% UI 4,500–27,000) due to non-AIDS cancers, childhood diarrhoea, maternal disorders, and neonatal disorders, reducing DALYs by 225,200, 609,600, 39,400, and 192,800 respectively (Supplementary Table S14). Adjusting for changes in population size reveals additional reductions in risk of kidney disease (7.5% reduction in DALYs per person-year), ALRI (6.8% reduction), and measles (8.9% reduction).

### 3.2 Healthcare services required for programme delivery

During 2010-2019, there were 433.8 million (433.3–435.5 million) appointments delivered for all health conditions including HTM services, encompassing community/outreach services, inpatient care, pharmacy and laboratory services, equivalent to 2.7 interactions per person per year (Figure 3). Through these appointments, the following HTM services were delivered: 157.0 million (95% UI 156.4–157.5 million) screening or diagnostic tests; 22.7 million (22.5– 22.9 million) preventive services including VMMC, IPT and PrEP; 23.2 million (23.1–23.2million) treatment and follow-up services; 558,700 (547,510–560,700) inpatient days were required.

**Figure 3:**
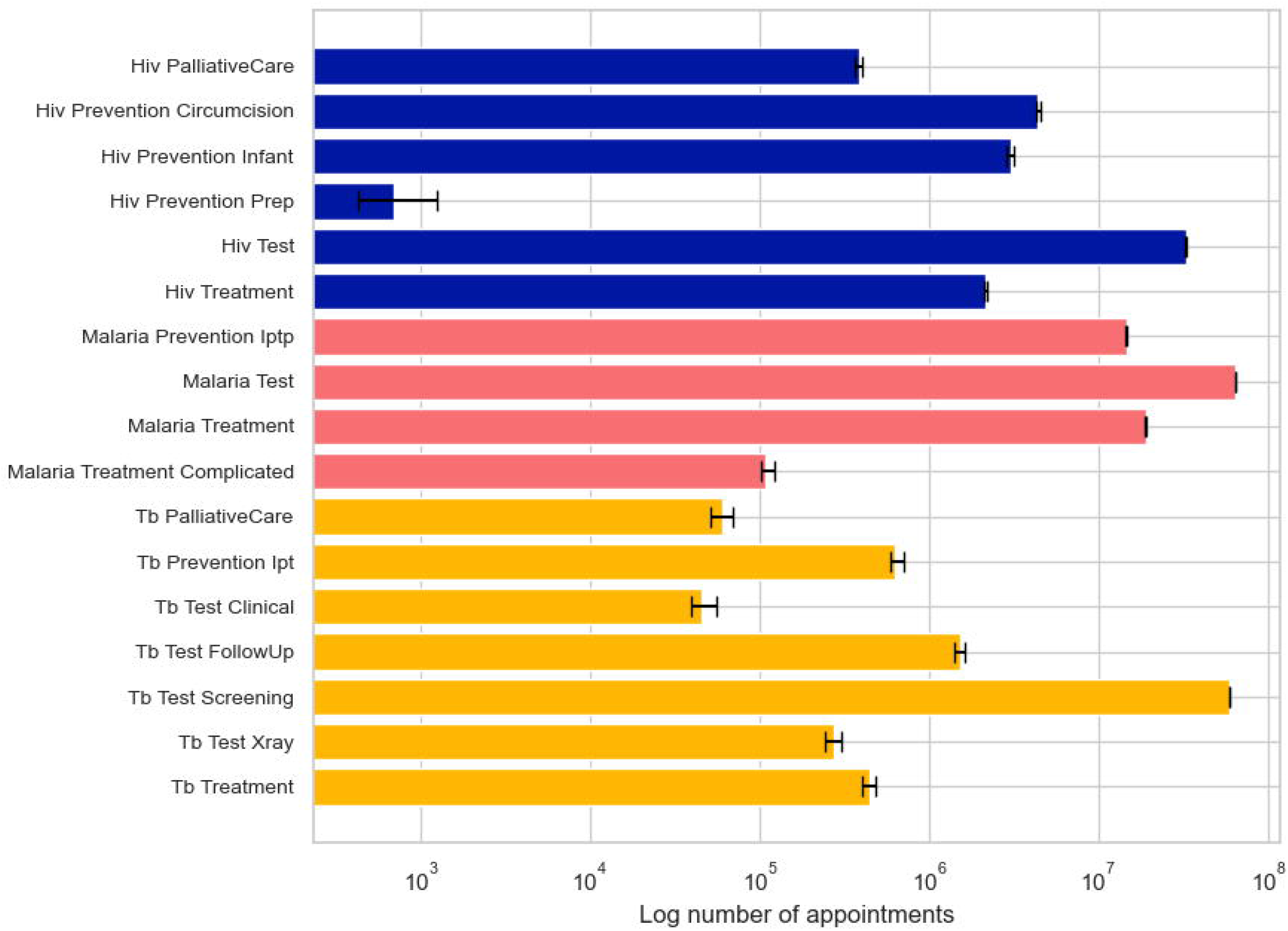
The numbers of HTM appointments estimated to have been delivered through each programme between 2010–2019. Some services are integrated within other appointments or delivered in the community.

From 2010–2019, HTM programmes accounted for 24.2% (24.1%–24.2%) of all healthcare services, decreasing from 22.3% (22.0–22.7%) in 2010 to 16.0% (15.3–16.2%) in 2019. Without these programmes, the apparent reduction in health system usage would be offset by the needs of untreated patients (Table 2). Specifically, there would have been 120.7 million fewer outpatient appointments, 2.0 million fewer laboratory services, and 323,500 fewer pharmacy visits, but 9.4 million additional hospital admissions and inpatient days, primarily for severe malaria and advanced HIV disease.

**Table 2:** Estimated hours of healthcare worker time (in millions) of each cadre required during the true provision of all healthcare services including HTM (Actual scenario) and if there had been no HTM services available. The laboratory services are currently only demanded in the model through the HIV, TB and malaria programmes.

| Cadre | Hours required under actual scenario | Hours required under no HTM scenario | Percentage change under no HTM scenario |
| --- | --- | --- | --- |
| Clinicians | 94.22<br>(93.24 - 94.53) | 81.94<br>(81.78 - 82.94) | -13.03% |
| Health Surveillance Assistants | 6.40<br>(6.38 - 6.45) | 0.05<br>(0.05 - 0.05) | -99.27% |
| Lab Technicians | 0.54<br>(0.53 - 0.55) | 0<br>(0 - 0) | -100.00% |
| Mental Health Workers | 0.07<br>(0.06 - 0.07) | 0.06<br>(0.06 - 0.07) | -3.49% |
| Nurses/Midwives | 113.74<br>(113.61 - 114.14) | 96.20<br>(96.11 - 97.08) | -15.42% |
| Pharmacists | 27.58<br>(27.42 - 27.69) | 22.82<br>(22.76 - 22.85) | -17.26% |
| Radiographers | 0.49<br>(0.48 - 0.49) | 0.41<br>(0.41 - 0.42) | -14.49% |
| **Total** | **243.04**<br>**(241.72 - 243.43)** | **201.49**<br>**(201.16 - 203.35)** | **-17.10%** |

The healthcare worker time required without HTM programmes would have been 201.5 million patient-facing hours, only slightly less than the 243.0 million hours actually used. Therefore, the adjusted cost in healthcare worker time required by HTM programmes over ten years was 41.5 million hours, or 17.1% of all patient-facing time. Most of this difference was due to reduced demands on Health Surveillance Assistants, who perform malaria tests and other community services.

## 4 Discussion

HTM programmes accounted for 24% of all health system interactions in Malawi from 2010 to 2019, resulting in significant health gains but demanding millions of hours of clinical, nurse and pharmacist time. HTM programmes have not only led to significant reductions in targeted diseases but have also reduced susceptibility to concurrent health challenges such as ALRI, diarrhoeal diseases, and neonatal disorders. In scenarios without HTM services, high early-life mortality rates generally lead to fewer non-communicable disease deaths later in life. Thus, estimating spillover effects of HTM services on other diseases provides a fair assessment of changes in disease burdens but may underestimate the impact on reducing per-capita mortality risks from other causes.

Empirical studies show significant life expectancy gains due to ART (10 year increase in adults) [40], malaria elimination (6 year increase in children) [41] and overall health improvements (10 year increase).[42] Our analysis demonstrates that the HTM programmes - comprising widespread ART access, enhanced TB control, and effective malaria prevention - implemented through all stages of life from pregnancy/delivery, through childhood and into adulthood, have collectively increased life expectancy from birth by 13.0 to 16.9 years and averted over 75 million DALYs within ten years.

In the absence of HTM services, the savings in health system demand are diminished due to the resulting deterioration in health. The substantial outpatient care required for HTM alleviates the burden on secondary and tertiary facilities, contributing to nearly 1 million fewer inpatient admissions (9.4 million bed-days) over the ten-year period. Healthcare workers face high TB rates due to limited infection control resources and over-crowded facilities.[43] Whilst we do not quantify the deaths averted in this population, protecting this population would further reduce health system pressure.

While this comprehensive framework serves as a good foundation for evaluating the collective impacts of diverse interventions on health and health systems, there are limitations as with any modelling approach. Despite the robust data available for HTM programmes, limitations in the data—such as gaps in key indicators and underreporting— persist. Expert input helps address these challenges, aligning outputs with existing models and programme data. Ongoing strategies, like active TB case finding through FAST, MDU screening of key and vulnerable populations, community interventions for HIV and TB testing and treatment adherence clubs continue to strengthen the programmes, leading to TB case incidence of 119 per 100,000 with 90% treatment success rates and approximately 11,000 AIDS deaths in 2023.[2, 8] The majority of laboratory technician and health surveillance assistant time in the model is currently utilised for HTM care, though these roles also encompass other services including routine diagnostics, disease monitoring, and emergency response, which are not fully captured in this model.

The model was designed to balance complexity and clarity by including key disease interactions while omitting others. Critical interactions are captured, but second-order effects, such as those between malaria and acute lower respiratory infections (ALRI), are excluded. Additionally, while malaria can cause vomiting and stomach aches, these symptoms are not directly linked to diarrhoeal pathogens in the model. HTM services provide cumulative benefits; for example, malaria interventions significantly impact under-five mortality rates 2.5 years after implementation.[44] The model uses a fixed 2018 estimate for consumables, which may not fully reflect changes from 2010 or evolving treatment guidelines. As a result, uncertainties in epidemiological and service delivery models could lead to overestimations of health impacts or healthcare worker time requirements.[45]

This study examines health outcomes and healthcare delivery, although HTM programmes also enhance infrastructure, workforce training, capacity building, and supply chain management.[46, 47]. Improved service delivery models have reduced the burden of HTM services on the health system while increasing health impact, indicating a move toward sustainability. The finding that 17% of healthcare worker time contributes to a more than 10-year increase in life expectancy reflects a highly favourable outcome. With greater efficiency, the overall health system effort could decrease further. As global health funding decisions are made, a holistic understanding of programme effectiveness and resource implications should guide prioritisation to maximise overall health impact.

## Data Availability

All data used are publicly available. The model source code and accompanying description are freely available on GitHub.

https://tlomodel.org

## Acknowledgements

We would like to express our gratitude to our colleagues at the National HIV, Viral Hepatitis & STI programme and the Department of Planning and Policy Development, Ministry of Health and Population, Malawi, for their continuous support of this project. We also wish to acknowledge the National AIDS Commission and the Health Economics and Policy Unit (HEPU) at Kamuzu University of Health Sciences for their valuable contributions and advice.

