## Supplementary Information for "A Decade of Progress in HIV, Malaria, and Tuberculosis Initiatives in Malawi"

### 1 Additional Methods

The methods detailed here can also be found in [1] and www.tlomodel.org. Further details on the health system interactions for HIV and TB care can also be found at these sources. All associated code is open-source and available on github (https://github.com/UCL/TLOmodel/releases/tag/tara_htm_scaleup_v1.0).

#### 1.1 HIV transmission model

##### 1.1.1 HIV transmission in adults

New HIV infections among individuals aged over 15 years are determined annually through a stochastic binomial selection process. The probability of infection is influenced by the transmission rate $\beta$ and the individual's susceptibility, which is shaped by their lifestyle attributes and demographic details (Table 1). The infection risk for women is influenced by the prevalence of non-virally-suppressed infections in men, and vice versa. We assume fully random sexual mixing, with an additional risk factor for female sex workers.

The infection risk for an individual $i$ with characteristics sex $s$, age $a$, and additional time-varying properties $m$ (residence urban/rural, wealth quintile, education level, exposure to behavior change intervention, pre-exposure prophylaxis, circumcision) at time $t$ is given by:

$$\lambda_i^{s,a,m}(t) = \frac{\beta}{N_s} \left[ \sum_{\hat{s},a \geq 15} I^{\hat{s},d}(t) \cdot h(\hat{s},a,d) \right] \cdot \mathrm{RR}_i^{s,a,m}(t) \tag{1}$$

where $I^{\hat{s},d}$ is the sum of infected individuals with sex $\hat{s}$ and treatment status $d$ which is multiplied by the relative infectiousness $h$ of each infected person. The relative infectiousness $h$ depends on the sex $\hat{s}$ of the infected person, and their age $a$ and treatment status $d$. The transmission rate $\beta$ is calibrated and fixed.

$$h(\hat{s},a,d) = \begin{cases} 1 & \text{if } s \neq \hat{s},\ 15 \leq a \leq 49, \text{ and } d = \text{ not treated or not virally suppressed} \\ 0 & \text{otherwise} \end{cases}$$

Treatment status $d$ can take on one of three values:

- **Untreated**: The individual has not received any treatment.
- **Treated but not virally suppressed**: The individual has received treatment, but the treatment has not fully suppressed the virus.
- **Treated and virally suppressed**: The individual has received treatment and the virus has been fully suppressed.

It is assumed that individuals who are **treated and virally suppressed** do not pose a risk of transmitting the infection to others. $N$ is the sum of susceptible (non-infected) individuals of sex $s$.

The individual relative risk $\mathrm{RR}_i^{s,m}$ at time $t$ is defined in Equation S3. The alpha terms here reflect the :

$$\begin{aligned}\mathrm{RR}_i^{s,m}(t) = \alpha_0 + \alpha_{\text{sex}} \cdot \mathrm{s}_i + \alpha_{\text{circ}} \cdot \text{Circumcision}_i(t) + \\ \alpha_{\text{PrEP}} \cdot \text{PrEP}_i(t) + \alpha_{\text{urban}} \cdot \text{Location}_i(t) + \alpha_{\text{wealth}} \cdot \text{Wealth}_i(t) + \\ \alpha_{\text{education}} \cdot \text{Education}_i(t) + \alpha_{\text{behaviour}} \cdot \text{Behaviour change counselling}_i(t)\end{aligned} \tag{2}$$

where:

- $\alpha_0$ represents the intercept, set to 1.
- $\alpha_{\text{sex}}$ quantifies the effect of the individual's sex on the risk of infection.
- $\alpha_{\text{circ}}$ reflects the impact of circumcision status on the risk of infection.
- $\alpha_{\text{PrEP}}$ accounts for the effect of current pre-exposure prophylaxis (PrEP) use on the risk of infection.
- $\alpha_{\text{urban}}$ represents the effect of living in an urban location versus a rural location on infection risk.
- $\alpha_{\text{wealth}}$ indicates the influence of wealth status on the infection risk.
- $\alpha_{\text{education}}$ measures the effect of education level on infection risk.

- $\alpha_{\text{behaviour}}$ represents the impact of exposure to behaviour change counselling on infection risk.

The HIV polling event runs each year and schedules new infections in adults aged over 15 years through horizontal transmission. The probabilities of transmission from males to females and vice versa are calculated separately. We do not differentiate between homosexual versus heterosexual transmission as data on the size of this population are scarce and we assume that intravenous drug use as a risk factor for HIV acquisition is not widespread in Malawi. The infection dates for all newly infected people are randomly distributed across the year.

###### 1.1.2 HIV transmission in children

Incident infections among individuals aged under 15 years result exclusively from mother-to-child transmission (see Figure 1). HIV transmission from an infected mother to the infant is stratified into two risk phases: gestational/delivery and breastfeeding and the hazard of infection to infants during pregnancy or delivery hinges upon the timing of maternal infection. If maternal infection occurs during pregnancy, the risk of mother-to-child transmission is higher than that associated with infections predating pregnancy (probability=0.3 if infection occurs during pregnancy, 0.22 if pre-pregnancy) (Rollins, Mahy et al. 2012). If infection does not occur in the infant during pregnancy or delivery, there remains a potential exposure to HIV transmission with ongoing breastfeeding. The time-frame for breastfeeding-induced infection is determined through a stochastic process involving a random draw from an exponential distribution with a rate parameter equivalent to the reciprocal of the monthly probability of mother-to-child transmission (monthly probability=0.01). If breastfeeding ceases before the scheduled infection event, the child remains HIV-negative.

If the mother is on antiretroviral therapy (ART) and virally-suppressed, vertical transmission risk is eliminated. Infants born to HIV-infected mothers on ART without viral suppression remain vulnerable to infection, similar to those born to untreated mothers. If mothers on ART are ART-adherent and virally-suppressed, the infant will concurrently receive zidovudine (AZT) or nevirapine (NVP) subject to availability until one week following the cessation of breastfeeding.

###### 1.1.3 Progression to AIDS and mortality

The expected survival time for infants infected before age 5 years either follows an exponential distribution (if infected prior to or during birth) or a Weibull distribution (if infected after birth), giving a median survival time of 0.64 years and 16 years respectively.([2]) The time from infection to AIDS is either equal to the survival time if less than 1 year or 18 months before the expected date of death. If infants are on ART and virally suppressed, AIDS onset and AIDS deaths will not occur.

For untreated (or treated but not virally-suppressed) adults, the expected survival time follows a 2-parameter Weibull distribution with shape parameter=2.55 and an age-dependent scale parameter calculated at the time of infection. The median survival time for men aged 25-34 is 10.6 years. The date of AIDS onset is randomly drawn from the exponential distribution with mean=18 months.

##### 1.2 TB fixed incidence model

TB is modelled through a fixed incidence model, adjusted by the prevalence of untreated active TB in the population. The WHO reported estimates of active TB incidence each year (in 2010, 338 active TB cases per 100,000 population, 95% CI 55 - 870) are adjusted by a calibrated scaling factor which converts reported incidence to a risk of active TB in the absence of any interventions, such as BCG and TB Preventive Therapy.([3]) The susceptible population is subject to this risk each year, weighted by their individual risk factors (Table 2) and infections are randomly distributed across the year. The susceptible population includes those with latent infections or untreated active infections. Each year, the proportion of multidrug-resistant TB (MDR-TB) strains is assumed constant at 1.86%. We define MDR-TB cases as infections which are resistant to either rifampicin treatment only or both isoniazid and rifampicin.

Infections due to drug-susceptible (DS) and MDR-TB are assigned separately, with the risk of applied to the population proportional to the current prevalence of untreated infections of that strain, including those with an MDR-TB infection who are currently on an ineffective first-line regimen. Individuals can be infected with either strain or concurrently with both, in which case clinical disease is assumed to arise from the most recent infection. We model here the incidence of active symptomatic TB directly from an uninfected (susceptible) stage. The person will progress to a latent stage of infection following self-cure or treatment, from which relapse or reinfection can occur.

The relative risk (RR) of infection (or reinfection) for individual $i$ with characteristics $m$ including age-group (divided into children under age 15 years and adults), BCG vaccination status, HIV infection (and ART use), lifestyle factors such as smoking or heavy alcohol use and whether the person is currently on TPT is calculated using a generalised linear regression model. The per capita risk of active TB infection for strain ($f \in \{ds, mdr\}$) is:

$$\text{Risk}_{\text{TB}_f}(t) = \text{scaling\_factor} \cdot \text{WHO\_incidence}(f) \cdot p(f) \cdot \text{RR}_{i,m}(t) \tag{3}$$

The proportion of untreated TB infections, denoted as $p(f)$, is calculated as the ratio of the sum of infectious individuals infected with strain $f$ who are untreated to the total number of infections of strain $f$:

$$p(f) = \frac{\sum_d (I(f,d) \cdot h(f,d))}{I(f)} \tag{4}$$

where $\sum_d (I(f,d) \cdot h(f,d))$ represents the sum of the number of infected individuals with strain $f$, each multiplied by their infectiousness $h(f,d)$, across all treatment statuses $d$. This sum is then divided by the total number of individuals infected with strain $f$, $I(f)$, to obtain the proportion of untreated infections.

The infectiousness $h(f,d)$ is averaged across smear-negative and smear-positive cases and is assumed to be equal among drug-sensitive and MDR-TB strains:

$$h(f,d) = \begin{cases} 1 & \text{if } f = \text{DS and } d = \text{no treatment,} \\ 1 & \text{if } f = \text{MDR and } d = \text{no treatment,} \\ 1 & \text{if } f = \text{MDR and } d = \text{non-MDR regimen,} \\ 0 & \text{otherwise.} \end{cases}$$

New active infections are assigned using a binomial draw with probability $\text{Risk}_{\text{TB}_f}$. Smear status and TB-symptoms (fever, respiratory symptoms, fatigue and night sweats) are determined at the onset of active TB infection. Individuals are assigned a smear status through a random draw with probability 0.62 (95% CI 0.42 – 0.80) in HIV-negative individuals and 0.35 (95% CI 0.19 – 0.54) in PLHIV.([4, 5, 6]). If a person is HIV+, the onset of active TB disease also indicates the onset of AIDS through the HIV module.

#### 1.3 Malaria emulation model

The risks of malaria are determined through an emulation model calibrated to age- and district-specific parasite prevalence data ([7]). This model generates lookup tables that assign risks of asymptomatic, clinical, and severe malaria for different age groups (infants aged 0-6 months, 6-12 months, and one-year age bands thereafter) by district and month. These tables account for various levels of bednet and indoor residual spraying coverage, with risks varying by district and month to reflect the geographical and seasonal distribution of malaria in Malawi.

For each month, the district-level incidence of infection, clinical malaria, and severe malaria are determined by referencing the appropriate values from the lookup tables, using the reported district-level estimates of bednet and indoor residual spraying coverage. These incidence rates are calculated for every age band.

For each susceptible individual (i.e., not currently experiencing clinical or severe malaria), their age band and individual characteristics are used to derive a probability of infection, calculated as the district-age incidence multiplied by the individual relative risk (Table 3). It is assumed that every individual is equally at risk of asymptomatic malaria infection, but the individual risk of clinical or severe malaria is influenced by factors such as HIV status, ART status (if virally suppressed), and preventive measures such as Intermittent Preventive Therapy for pregnant women and Cotrimoxazole.

Among those infected, individuals with asymptomatic malaria are assessed for the risk of progressing to clinical infection using the lookup table values for clinical incidence by age band and individual risk factors. A similar process is used to identify new severe cases from the pool of clinical cases. Here, a random draw determines the transition to severe malaria, based on probabilities derived from the lookup tables and individual characteristics.

Each month, this random draw assigns all new malaria infection types and distributes the onset of infection or disease throughout the month. Upon onset of clinical malaria, a set of symptoms (fever, headache, vomiting, stomach ache) is applied to each affected individual, potentially prompting healthcare-seeking behaviour through an algorithm that considers the presence of symptoms along with individual characteristics such as age, education, and wealth. Pregnant individuals will additionally experience severe anaemia, and severe malaria cases will require emergency care.

Clinical malaria and the associated symptoms are assumed to last for five days, resolving either through treatment or natural recovery. Parasitaemia is cleared by treatment within 21 days of treatment initiation. If untreated, detectable parasitaemia persists for 195 days in clinical cases and 110 days in asymptomatic cases ([8, 9, 10]). Severe malaria symptoms and parasitaemia can only be resolved through appropriate treatment.

##### 1.3.1 Malaria interventions

*Malaria rapid diagnostic test (RDT)*

The National Malaria Policy in Malawi recommends the use of RDTs across all facilities with microscopy testing reserved for severe malaria cases. The majority of RDTs used across sub-Saharan Africa target P. falciparum histidine-rich protein 2 PfHRP2 in the peripheral blood, however the clinical sensitivity of these tests depends on the population parasite density which in is driven by the transmission intensity.

Studies in Malawi have shown RDT sensitivity varying between 90-92% depending on the manufacturer, with lower specificity (39-68%).([11]) A broader meta-analysis reported higher sensitivity (95.0%, 95% CI 93.5-96.2%) and specificity (95.2%, 93.4-99.4%) for PfHRP2 RDTs in endemic areas. However, these tests can also detect recently treated or self-resolved infections for several weeks.([12]) For this analysis, we assume a conservative sensitivity of 95% and a specificity of 100% for RDTs in detecting malaria parasitaemia.

If RDTs are not available at the time of the appointment, a person can return to a health facility in their district up to 5 times before defaulting from care.

*Malaria treatment*

The consumables required for the provision of malaria treatment are detailed in Table 4. If the required treatment is not available at a health facility on the day of the appointment, a person can return up to five times before defaulting from care. For severe (complicated) malaria cases, a person can be admitted to a general hospital bed even if malaria treatment is not available, in which case they would receive any consumables from the package of care which are available, e.g. paracetamol.

*Intermittent preventive therapy for pregnant women*

Each dose of Sulfadoxine / Pyrimethamine (SP) used for intermittent preventive therapy for pregnant women clears both asymptomatic and symptomatic infections and provides up to six weeks of post-treatment prophylaxis preventing further infection.(White 2005) Pregnant women in Malawi are recommended to receive at least three doses of SP given four weeks apart after the first trimester of pregnancy during each scheduled antenatal care visit. SP is contraindicated for women receiving Cotrimoxazole. The reductions in risks of clinical and severe malaria in Table 3 are applied after two doses of SP and protection wanes after six weeks.

Pregnant women with HIV are recommended to use Cotrimoxazole in place of IPTp daily and throughout the duration of the pregnancy.

*Cotrimoxazole*

Individuals infected with HIV are routinely prescribed Cotrimoxazole alongside ART. We assume that the protective effects of Cotrimoxazole applies equally to incidence of both clinical and severe malaria and are consistent across all age-groups.

#### 1.4 DALY weights

The aggregate impact of all causes of mortality and morbidity in the population is summarised using Disability-Adjusted Life Years (DALYs), which combine Years of Life Lost (YLL) due to premature death and Years Lived with Disability (YLD) due to illness. The total DALYs are calculated as DALY=YLL+YLD. YLL is recorded when an individual dies before age 70, attributing each subsequent lost day to the cause of death. YLD is recorded monthly, reflecting the average disability burden of each disease on each individual, with adjustments made for comorbidities. The disability weights are sourced from the Global Burden of Disease Study and are recorded for each individual in the simulated population by time period, age, and cause.[13]

#### 1.5 Disease interactions

In this section we detail the disease interactions associated with HIV, TB and malaria infections, although many other interactions between health conditions are captured in the Thanzi la Onse model. This feature of the model has been adapted from [1] and extended to include a number of non-communicable diseases, including mental health. The interactions associated with HIV, TB and malaria are detailed below:

##### 1.5.1 Interactions with HIV

**Tuberculosis**

- Individuals with untreated (or treated but not virally suppressed) HIV are at a higher risk of developing active TB infection.
- Individuals with untreated (or treated but not virally suppressed) HIV who contract active TB have an increased likelihood of developing smear-negative TB.
- Concurrent active TB and HIV infection automatically classify an individual as having AIDS, thus increasing the risk of AIDS-related mortality.
- Individuals with HIV who are not virally suppressed cannot clear an active TB infection without treatment; self-cure of TB is only possible in those who are HIV-negative or HIV-positive and virally suppressed.
- The risk of TB relapse is higher among individuals with HIV.

**Malaria**

- Individuals with untreated (or treated but not virally suppressed) HIV are at a higher risk of developing clinical and severe malaria.
- Pregnant women with untreated (or treated but not virally suppressed) HIV have higher risks of clinical and severe malaria.

**Pneumonia**

- Individuals with HIV (whether untreated or treated but not virally suppressed) face higher risks of developing acute lower respiratory infections (ALRI). This increased risk applies uniformly across all causative agents of ALRI. The pathogens associated with ALRI include:
  - Respiratory Syncytial Virus
  - Rhinovirus, Human Metapneumovirus
  - Parainfluenza Virus
  - Streptococcus pneumoniae (including Pneumococcal Conjugate Vaccine (PCV) 13 serotypes and non-PCV13 serotypes)
  - Haemophilus influenzae (both type B and non-type B)
  - Staphylococcus aureus
  - Enterobacteriaceae (including E. coli and Klebsiella)
  - other Streptococci or Enterococci
  - Influenza Virus
  - Pneumocystis jirovecii
  - other viral pathogens
  - other bacterial pathogens, and unspecified pathogens.
- Individuals with untreated (or treated but not virally suppressed) HIV have a higher risk of treatment failure for ALRI.

**Diarrhoea**

- Children with untreated (or treated but not virally suppressed) HIV are at a higher risk of developing childhood diarrhoea caused by various pathogens, including:
  - Rotavirus
  - Shigella
  - Adenovirus
  - Cryptosporidium
  - Campylobacter
  - Enterotoxigenic Escherichia coli (ETEC)
  - Sapovirus
  - Norovirus
  - Astrovirus
  - Typical Enteropathogenic Escherichia coli (tEPEC)
- Individuals infected with one of these diarrhoeal pathogens and with untreated HIV will have prolonged symptom duration
- Mortality rates attributed to diarrhoeal pathogens will be higher in individuals co-infected with HIV and not virally suppressed, compared to those without HIV infection.

**Schistosomiasis**

- Women with high Schistosoma haematobium worm burdens are at increased risk of HIV infection acquisition.

**Anaemia**

- Pregnant women with untreated (or treated but not virally suppressed) HIV have higher risks of anaemia during pregnancy (independent of parity) and post-pregnancy up to week 6 of the postnatal period.

**Stunting**

- Children aged under 5 years with untreated (or treated but not virally suppressed) HIV experience higher incidence and severity of stunting.

**Depression**

- Individuals with untreated (or treated but not virally suppressed) HIV have higher risks of onset of depressive disorder compared with the HIV-uninfected population.

**Cardiovascular disease**

- The risks of cardiovascular disease, including myocardial infarction, ischaemic heart disease, stroke or coronary heart disease are higher in people with untreated (or non-virally-suppressed) HIV infection.

**Non-AIDS cancers**

- Incidence of non-AIDS cancers is increased in individuals with untreated (or treated but not virally suppressed) HIV.

##### 1.5.2 Interactions with TB

**Diabetes**

- Risk of active TB is increased in people with untreated Type I Diabetes Mellitus.
- Risk of death due to TB is increased in people with untreated Type I Diabetes Mellitus.
- Risk of TB relapse is increased in people with untreated Type I Diabetes Mellitus.
- All of the effects above are adjusted for HIV, age and sex as counfounders.

##### 1.5.3 Interactions with Malaria

**Preterm birth**

- The risk of preterm labour is higher in women with malaria parasitaemia, independent of level of parasitaemia and/or presence of symptoms. This risk is not impacted by parity. In turn, preterm labour increases the risk of childhood stunting.

**Maternal anaemia**

- The risk of anaemia during pregnancy and during the post-partum period is higher in women with malaria parasitaemia, independent of level of parasitaemia and/or presence of symptoms. This risk is not impacted by parity.

**Stillbirth**

- The risk of stillbirth is higher in women with malaria parasitaemia, independent of level of parasitaemia and/or presence of symptoms. This risk is not impacted by parity.

#### 1.6 Integrated health care

As detailed above, many individuals will experience multiple, concurrent health challenges and can seek care for specific symptoms or health conditions. The Malawi Standard Treatment Guidelines indicate multiple tests can be delivered within appointments and certain symptoms can trigger referrals for alternative diagnostic tests or treatment plans. These will differ for the individual circumstances of each patient. Some examples of this include:

- HIV tests are offered to all individuals undergoing TB screening
- Any HIV diagnosis will automatically trigger TB screening for clinical symptoms. Where symptoms are indicative of an active TB infection, a referral for a TB diagnostic appointment will occur
- Infants born to mothers with diagnosed HIV infection will have two HIV tests offered within the first few months of life. Infants who remain HIV-negative are given prophylaxis throughout the breastfeeding period
- All Antiretroviral Therapy dispensations include Isoniazid Preventive Therapy (TPT) and Cotrimoxazole to protect against active TB and clinical/severe malaria respectively
- The choice of TB diagnostic test will depend on current HIV status; people diagnosed with HIV will have Gene Xpert recommended as a first-line TB diagnostic test
- Routine antenatal care services include HIV tests, distribution of insecticide-treated bednets and Intermittent preventive therapy for pregnant women

#### 1.7 Estimating life expectancy

We developed life tables for the simulated model population by calculating age-specific and sex-specific deaths rates for people age 0, 1-4, and in 5-year age-bands up to age 90+ thereafter. Model life tables were developed for males and females separately for each scenario using standard life-table methods.[14] The mortality rates at age $x$ are derived from the simulation outputs using the number of deaths in each age-sex band during 2019 divided by the population size in that band. The life expectancy of a person at birth is given by:

$$e_x = \frac{T_x}{I_x} \tag{5}$$

where $T_x$ is the total number of person-years lived by the cohort from age $x$=*0* until all cohort members have died and $I_x$ is the survivorship function, i.e. the number of persons in the cohort alive at age $x$=*0*.

#### 1.8 Health worker time

Healthcare services are delivered during health facility appointments, which may entail multiple services for different health conditions. These services occur at a particular facility level, requiring a fixed amount of healthcare worker time. We follow Berman *et al* [15] in using 50 different appointment types, and classify all healthcare services under one of these appointment types. The full list of appointment types and their healthcare worker time requirements are listed on the TLO Model website (www.tlomodel.org) and those services relating to HTM care are detailed below. If there is no specific appointment footprint relating to a particular HTM service, we state in the notes how we calculate the time requirement for service delivery.

Length of inpatient stay for TB palliative care is estimated from [16] and based on estimates of hospital stay across all TB cases (including those with subsequent treatment success). We use the maximum hospital stay across all TB cases as the average length of stay for an end-stage complex TB case with treatment failure.

#### 1.9 Model simulations

The model is an individual-based simulation implemented in the Python programming language, using the pandas data analysis library.[17] Each modelled process—encompassing disease, demographic, and behavioural factors—is encapsulated within a code module that contributes ’events’ to a queue. The simulation executes these events in chronological order, with events accessing and modifying a centralised store of individual characteristics. Healthcare service delivery is a specialised category of events that are subject to checks on required healthcare resources.

The simulation is initialised on January 1, 2010, with a representative model population scaled to 1:100. The median estimates and 95% uncertainty ranges of each output are derived across five model runs, initialising with identical starting parameters but deviating through stochastic (random) processes which incorporate the uncertainty around many parameter values.

The details of all demographic and disease-related events, as well as healthcare service delivery events are compiled and logged at the end of each year. The Disability-Adjusted Life Years (DALYs) incurred in the population are computed as the sum of Years of Life Lost (YLL) and Years Lived with Disability (YLD). YLL is calculated as the difference between the age at death and a reference life expectancy value. YLD is calculated using disability weights for each health condition, as provided by Salomon et al.[13] The disability for each individual, from all conditions, is measured monthly and combined additively.

### 2 Section 2: Additional Results

#### 2.1 Epidemiological outputs

The epidemiological outputs between 2010 and 2010 (incidence per 1000 person-years and mortality rates per 1000 person-years) are shown in 3.

#### 2.2 Full breakdown of deaths incurred

The summary of deaths due to HIV, TB, malaria and all causes with the associated estimated life expectancy is shown in Table 9. Full breakdown of the estimated mortality rates by each cause is represented in Table 10.

#### 2.3 Estimated deaths by age-group

Figure 6 shows the numbers of deaths occurring in each simulated scenario disaggregated by age-group.

#### 2.4 Full breakdown of DALYs incurred

Tables 12, 13 and 14 show the full breakdown of DALYs by cause during 2010 - 2019 through each of the scenario simulations. These values represent the median across the 5 runs of each draw and the 95% uncertainty intervals.

#### 2.5 Health system usage during 2010-2019

The full breakdown of appointment types estimated to have been delivered through routine health services and in the hypothetical scenario excluding HTM services is detailed in 15.

#### 2.6 Service provision required to deliver HTM services

HTM services can be aggregated into four distinct service types (prevention, testing, treatment, inpatient care) as detailed below. The median number of healthcare service classified into these groups along with 95% uncertainty intervals are calculated across the five runs of the Actual scenario (16).

**Preventive** = HIV infant prophylaxis, voluntary medical male circumcision (VMMV), HIV pre-exposure prophylaxis (PrEP), TB Preventive Therapy (TPT), intermittent preventive treatment of malaria during pregnancy
**Testing** = HIV test, TB screening, TB sputum test, TB GeneXpert test, TB chest x-ray, malaria test (mRDT)
**Treatment** = HIV treatment, TB treatment, TB follow-up, malaria treatment
**Inpatient** = HIV palliative care, TB palliative care, malaria treatment (complicated)

The estimated time requirements to deliver healthcare services are determined using the appointment footprint for every service, averaged across facility levels. For example, malaria treatment for a child aged under 5 years will require an "Under5OPD" appointment, which uses 20.9 minutes of clinical worker time, 12.7 minutes of nursing time and 5.4 minutes of pharmacist time.

The broad appointment types are further categorised into four groups, outpatient, inpatient, laboratory services and pharmacy. The mapping of each appointment type to a category is below:

**Outpatient** = Voluntary medical male circumcision, family planning, voluntary testing and counselling for HIV-negative person, outpatient appointment for person aged over 5 years, voluntary testing and counselling for HIV-positive person, minor surgery, outpatient appointment for person aged under 5 years, HIV-related appointment for new ART initiation, paediatric appointment for HIV services, routine appointment for established medically non-complex HIV patient, immunisation, under 5 malnutrition, consultation with health surveillance assistant, newly diagnosed TB patient, mental health outpatient appointment, follow-up for TB patient, first antenatal appointment, subsequent antenatal appointment, uncomplicated labour and delivery
**Inpatient** = inpatient admission, inpatient bed-day, major surgery, complicated labour and delivery, Caesarean

section, accident and emergency
**Laboratory** = diagnostic radiography, tomography, mammography, TB microscopy, molecular diagnostics
**Pharmacy** = pharmacy dispensing

### 3 Sensitivity analysis: estimating the impact of single programmes

When exploring the impact of each programme in the main analyses, we define the HIV-only impact as the difference between the full HTM programme and a hypothetical scenario whereby HIV services are excluded:

HIV programme impact = HTM services impact (Actual) - TB and Malaria service alone (exclude HIV services)

This contrasts with the 'siloed' approach adopted by other models, where the calculation is:

HIV programme impact = HIV programme impact - Impact of no HTM services

Here, siloed models measure programme impact against a baseline scenario where no HTM services are available, whereas our method evaluates the impact of fully integrated programmes versus scenarios where each component is hypothetically removed.

In our integrated approach, individuals connected to both HIV and TB programmes benefit from either intervention - those not saved by one are potentially saved by the other. Conversely, in a siloed model, such individuals could be counted twice - once under each programme - leading to double-counting.

To further investigate this, we conducted three additional scenarios:

1. **Exclude Malaria and TB**: Provides an estimate of the HIV programme's impact.
2. **Exclude HIV and Malaria**: Provides an estimate of the TB programme's impact.
3. **Exclude HIV and TB**: Provides an estimate of the malaria programme's impact.

The numbers of deaths, mortality rates and estimated life expectancy under these additional scenarios are presented in Table S17. Table S18 presents the estimated deaths averted by HIV, TB, and malaria (HTM) programmes using an alternative approach compared to the main analysis. This method involves evaluating the impact of each programme individually versus having no HTM services. The deaths averted are calculated separately for each disease: the number of deaths averted by the HIV programme (HIV/AIDS deaths), the TB programme (TB deaths), and the malaria programme (malaria deaths). The total HTM deaths averted is the sum of these individual contributions, compared against a scenario with no HTM services, as detailed in the main text.

Interestingly, estimating the deaths averted by each programme separately underestimates the total deaths averted compared to a joint estimation, contrary to initial expectations. This discrepancy is evident in the estimates of HIV/AIDS deaths averted and may be attributed to the interactions captured within the model. In the main analysis, when both TB and malaria programmes are operational, an additional 100,000 HIV/AIDS deaths are averted, suggesting that TB and malaria programmes indirectly save lives by preventing HIV/AIDS deaths.

Examining individual programmes reveals that focusing solely on HIV programmes leads to an increase in TB and malaria deaths due to reduced mortality from HIV, resulting in longer exposure to TB and malaria. Similarly, the TB programme alone results in higher HIV deaths, and the malaria programme alone increases HIV deaths by saving lives early in childhood, thus extending the period at risk for HIV infection.

In scenarios where all three programmes operate jointly, the synergistic effects are evident, as one intervention can prevent a death from one disease and later prevent death from another. For instance, preventing a malaria death can later prevent an HIV death. Conversely, in the absence of HIV services, high HIV mortality results in fewer individuals living long enough to be infected by malaria.

The analysis shows that jointly, the HTM programmes avert approximately 1.1 million deaths, demonstrating that the combined effect of the programmes is greater than the sum of their individual impacts. Mortality rates per 1,000 person-years show a greater reduction for the combined HTM programs compared to individual programmes: reduction of 6.97 deaths per 1,000 person-years for combined programmes versus 6.25 deaths per 1,000 person-years for single programmes. This suggests a synergistic effect where multiple programmes targeting different diseases amplify each other's impact, resulting in a larger overall reduction in mortality.

### 4 Figures and Tables

#### List of Figures

1 Schematic representing the risks of mother-to-child transmission for infants born to mothers with virally-unsuppressed HIV. . . . . 15
2 Schematic representing the structure of the malaria natural history model. . . . . 16
3 The annual incidence per 1000 person-years (upper figures) and mortality rates per 1000 person-years (lower figures) for HIV/AIDS, TB and malaria estimated under each scenario. The solid lines show the median estimates of five runs and the 95% uncertainty intervals are represented by the shaded areas. 16
4 Population pyramid depicting the true size of the population in 2019 by age-group (Actual) compared with the hypothetical scenario had no HTM services been available. . . . . 17
5 The true population size (Actual) over time compared with the hypothetical population size had HTM services not been available. Each line represents the output from one run. . . . . 18
6 The distribution of deaths by age-group occurring during the simulation period 2010 - 2019 by scenario. The bar labels "No ..." refer to the exclusion of services directly targeting that disease or diseases. Age_Grp: age-group . . . . . 19

#### List of Tables

1 Relative risk associated with acquisition of new HIV infection. The relative risks for sex, location and education were derived through regression modelling of DHS data 2015-2016. Relative risk for women engaged in sex work was derived through calibration, fitting the prevalence of HIV in female sex workers to data.([18, 19]) Relative risk for circumcised men was taken from ([20]) and the relative risk for those exposed at some point in their lifetime to behaviour change counselling is assumed. . . . . 20
2 Relative risks associated with active TB infection included in the TLO model. Abbreviations: TPT, Tuberulosis Preventive Therapy, which comprised mostly Isoniazid Preventive Therapy between 2010-2010; BMI, body mass index. . . . . 21
3 Relative risks of HIV and key interventions associated with clinical and severe malaria infections. . . . 21
4 Treatment options for malaria following the Malawi Standard Treatment Guidelines 2015 ([40]) . . . 22
5 DALY weights associated with HIV, TB and malaria infections. DALY weights for drug-susceptible and multidrug-resistant TB strains are assumed to be identical.[13] . . . . . 23
6 Key interactions between HIV, TB, malaria, and other disorders included in the TLO model. RR = relative risk. . . . . 24
7 The patient-facing time required for specific healthcare services in minutes. The patient-facing time is an example of the time taken for a standalone appointment to deliver each service. In practice, and in the simulation, these services may be integrated with other services, reducing the human resources requirement. Abbreviations: Over5OPD - patient over 5 years outpatient appointment; VCTNeg - voluntary testing and counselling for HIV-negative person; Peds - paediatric appointment; VCTPos - voluntary testing and counselling for HIV-positive person; EstNonCom - established non-medically-complex HIV patient; IPAdmission - inpatient admission. . . . . 25
8 The annual age-specific mortality rates estimated using the simulated numbers of deaths per population in 2019. Outputs are shown from one run. . . . . 25
9 Total deaths (thousands) estimated under each scenario and estimated life expectancy from birth for males and females using the all-cause mortality rates in 2019. . . . . 26
10 Median mortality rates per 1000 person-years with 95% uncertainty intervals by scenario. Mortality rates are calculated as the numbers of cause-specific deaths divided by person-years for each run and then aggregated by scenario. . . . . 26
11 Total deaths averted due by HIV/AIDS, TB and malaria cause rounded to the nearest 100. Single programme estimates are shown for the hypothetical single programme exclusion scenarios (e.g. HIV/AIDS deaths estimated had no HIV services been available, TB deaths had no TB services been available) and then calculated jointly had no HTM services been available. . . . . 26
12 Percentage change in DALYs incurred per person-year (median, lower and upper 95% uncertainty intervals) estimated when no HTM services are available compared with the Actual scenario. . . . . 27
13 Total DALYs incurred grouped by AIDS, TB (non-AIDS), malaria and other causes (non-HTM) during the simulation divided by person-years. Values are summarised across the runs of each draw. . . . . 27
14 Median DALYs incurred (thousands) with 95% uncertainty intervals by scenario. . . . . 28

15 The numbers of appointments (in thousands) classified by disease programme for the Actual scenario and if no HTM services had been available. The remaining HIV, TB and malaria services in the "No HTM" results relate to end-of-life or palliative care. First Attendance appointments refer to general outpatient appointments which are stratified into emergency and non-emergency appointments. The median values of 5 runs are shown for each scenario along with the lower (2.5th percentile) and upper (97.5th percentile) bounds. . . . 29
16 Estimated numbers of health system services required in millions for the provision of HTM care. . . . 29
17 Numbers of deaths (thousands) due to HIV/AIDS, TB, malaria and all causes; mortality rates due to HIV/AIDS, TB, malaria and HTM combined per 1000 person-years; life expectancy from birth estimated under the additional scenarios. Life expectancy for males and females is calculated using the all-cause mortality rates in 2019. . . . 30
18 Numbers of deaths averted (thousands) due to the HIV/AIDS, TB, malaria programmes separately and when no HTM services are available (main analysis). The estimate of HTM deaths averted using the single programme estimates is the sum of the HIV/AIDS, TB and malaria deaths averted through the single programmes (highlighted in blue). . . . 30

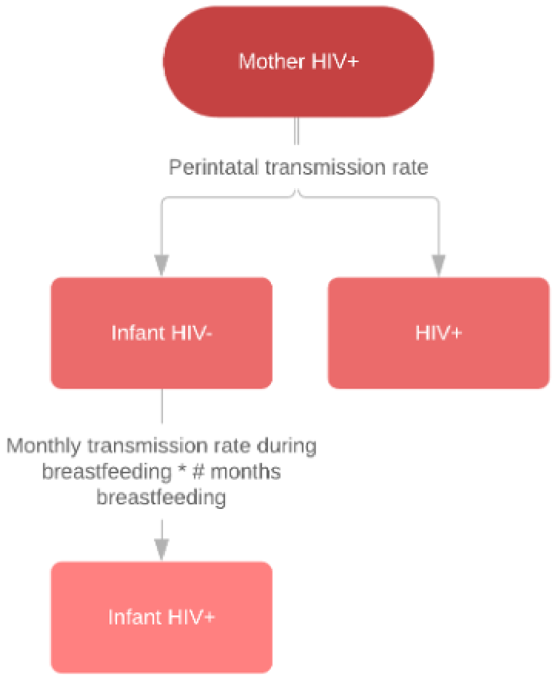


Figure 1: Schematic representing the risks of mother-to-child transmission for infants born to mothers with virally-unsuppressed HIV.

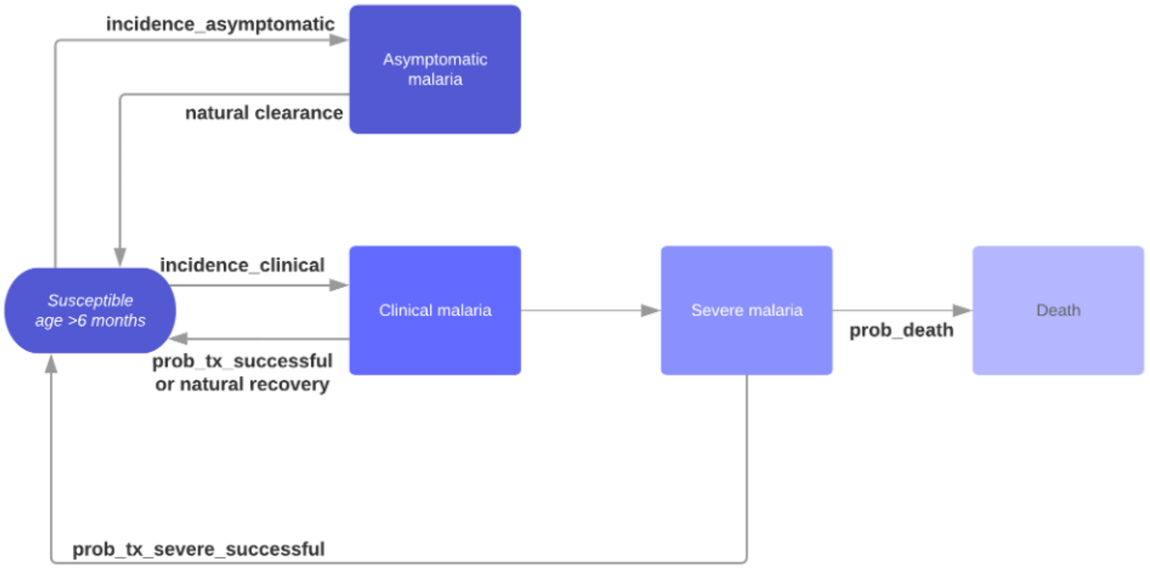


Figure 2: Schematic representing the structure of the malaria natural history model.

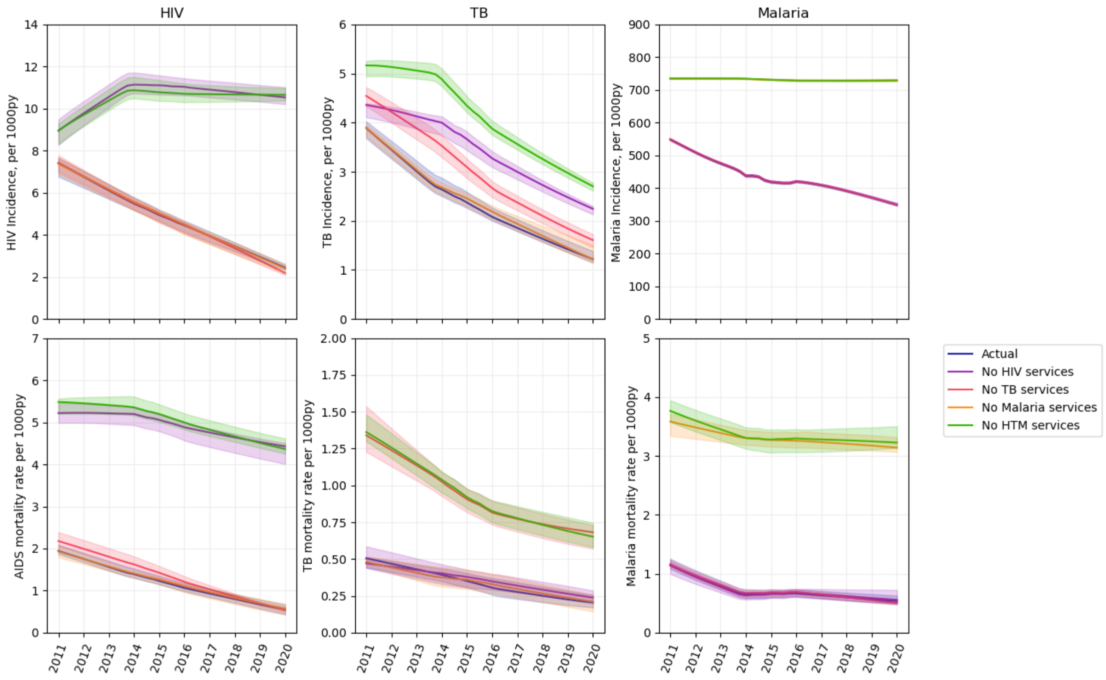


Figure 3: The annual incidence per 1000 person-years (upper figures) and mortality rates per 1000 person-years (lower figures) for HIV/AIDS, TB and malaria estimated under each scenario. The solid lines show the median estimates of five runs and the 95% uncertainty intervals are represented by the shaded areas.

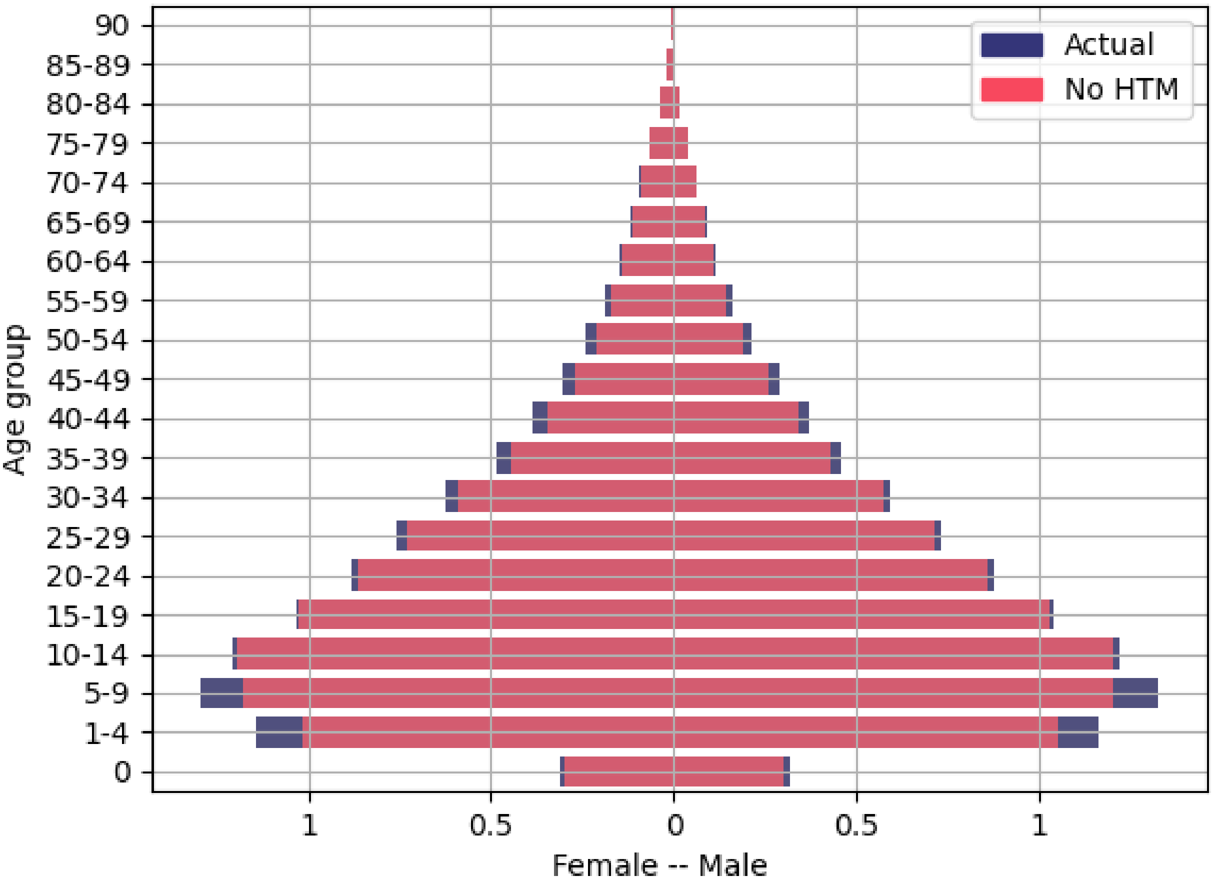


Figure 4: Population pyramid depicting the true size of the population in 2019 by age-group (Actual) compared with the hypothetical scenario had no HTM services been available.

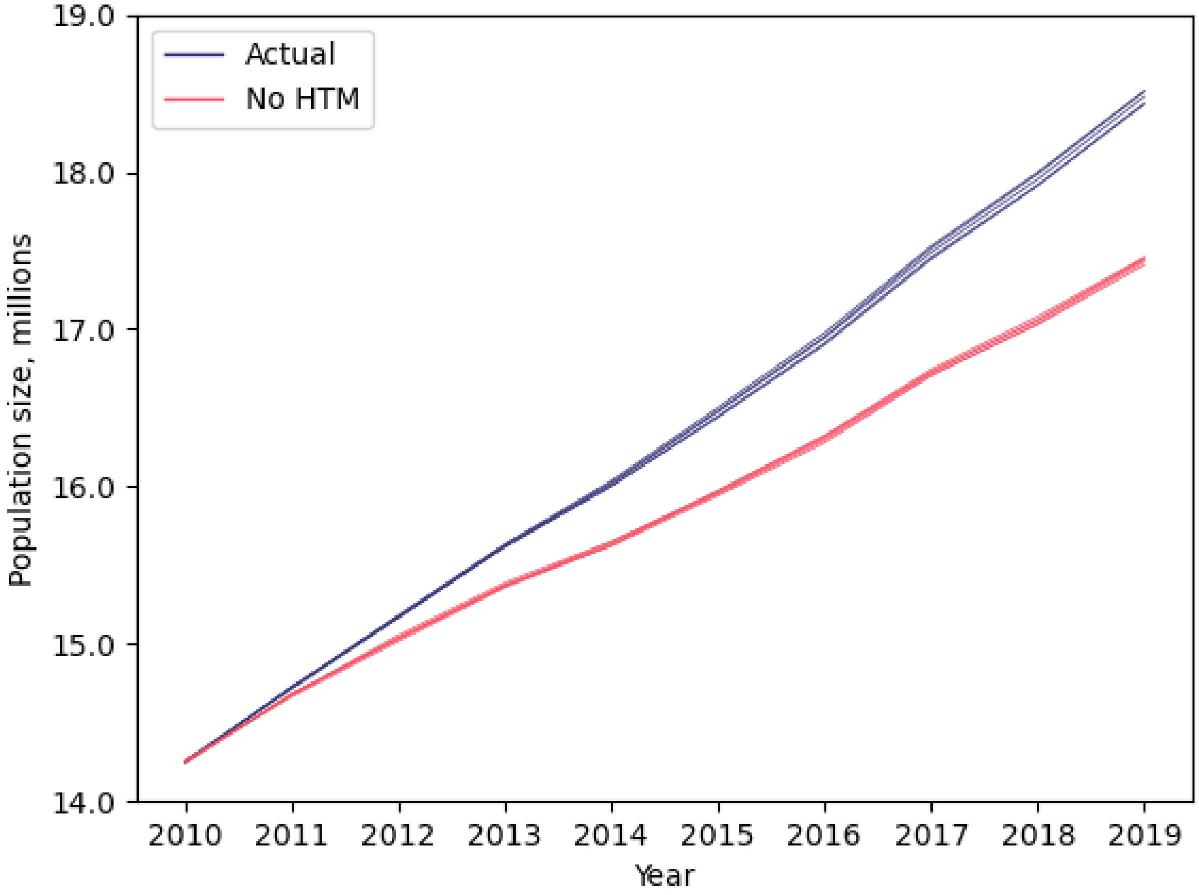


Figure 5: The true population size (Actual) over time compared with the hypothetical population size had HTM services not been available. Each line represents the output from one run.

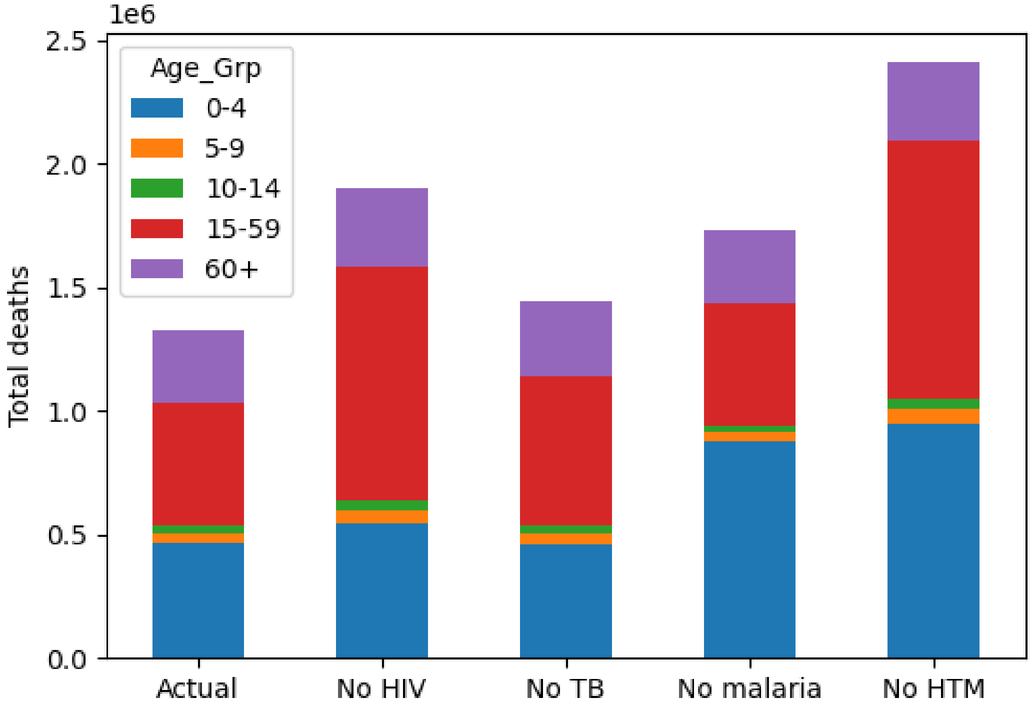


Figure 6: The distribution of deaths by age-group occurring during the simulation period 2010 - 2019 by scenario. The bar labels "No ..." refer to the exclusion of services directly targeting that disease or diseases. Age_Grp: age-group

| **Individual characteristic** | **Relative risk** |
|---|---|
| Sex | |
| Male | 1.0 |
| Female | 1.43 |
| Circumcision | |
| No | 1.0 |
| Yes | 0.4 |
| Pre-exposure prophylaxis | |
| No | 1.0 |
| Yes | 0.1 |
| Location | |
| Urban | 1.0 |
| Rural | 0.52 |
| Wealth quintile | |
| Poorest | 1.0 |
| Poorer | 0.96 |
| Middle | 1.18 |
| Richer | 1.19 |
| Richest | 1.56 |
| Education quartile | |
| None | 1.0 |
| Primary | 1.19 |
| Secondary | 1.15 |
| Higher | 1.0 |
| Behaviour change counselling | |
| No | 1.0 |
| Yes | 0.75 |
| Female sex work | |
| No | 1.0 |
| Yes | 25 |

Table 1: Relative risk associated with acquisition of new HIV infection. The relative risks for sex, location and education were derived through regression modelling of DHS data 2015-2016. Relative risk for women engaged in sex work was derived through calibration, fitting the prevalence of HIV in female sex workers to data.([18, 19]) Relative risk for circumcised men was taken from ([20]) and the relative risk for those exposed at some point in their lifetime to behaviour change counselling is assumed.

| Status | Relative risk of active TB | Source |
|---|---|---|
| Age in years | | |
| Adult $\geq 15$ | 1.0 | |
| Child <15 | 0.1 | Assumed, given proportional prevalence in children versus adults |
| BCG | 0.81 | [21] |
| TPT | | |
| Child HIV- | 0.55 [0.40-0.75] | [22] |
| Adult HIV- | 0.4 | [23, 24] |
| Child HIV+ | 0.31 [0.11-0.87] | [25] |
| Adult HIV+ | 0.68 [0.54-0.85] | [26] |
| Body mass index | | |
| BMI <30 | 1.0 | |
| BMI $\geq$ 30 | 0.4 | [27] |
| Heavy alcohol use ($\geq$40g per day) | 2.9 | [28] |
| Type 1 diabetes | 3.11 | [29] |
| Smoking status | | |
| Non-smoking | 1.0 | |
| Current smoking | 1.3 | [30] |
| HIV status | | |
| HIV- | 1.0 | |
| HIV+ (pre-AIDS) | 13 | Assumed to be half the value of HIV+ with AIDS |
| HIV+ with AIDS | 26.06 | [31] |
| On ART and virally suppressed | | |
| Child | 0.30 [0.21-0.39] | [32] |
| Adult | 0.35 [0.28-0.44] | [33] |

Table 2: Relative risks associated with active TB infection included in the TLO model. Abbreviations: TPT, Tuberculosis Preventive Therapy, which comprised mostly Isoniazid Preventive Therapy between 2010-2010; BMI, body mass index.

| Individual characteristic | Relative risk | Source |
|---|---|---|
| Intermittent preventive therapy for pregnant women | 0.54 (clinical)<br>0.6 (severe) | [34] |
| HIV+ and age under 5 years | 1.7 (clinical)<br>9.69 (severe) | [35, 36] |
| HIV+ and age over 5 years | 2.6 | [35] |
| HIV+ and pregnant | 3.96 (clinical)<br>2.8 (severe) | [37, 38] |
| Antiretroviral therapy (if HIV+) | 0.74 | [39] |
| Cotrimoxazole | 0.58 | [39] |

Table 3: Relative risks of HIV and key interventions associated with clinical and severe malaria infections.

| Age | Treatment |
|---|---|
| **Uncomplicated – 1st line** | |
| Adults | Malaria test kit (RDT)<br>Lumefantrine 120mg/Artemether 20mg |
| Children 0-5 years | Paracetamol 500mg<br>Malaria test kit (RDT)<br>Lumefantrine 120mg/Artemether 20mg |
| Children 5-15 years | Paracetamol syrup 120mg/5ml<br>Malaria test kit (RDT)<br>Lumefantrine 120mg/Artemether 20mg |
| First trimester - uncomplicated | Paracetamol syrup 120mg/5ml<br>Quinine sulphate 300mg<br>Clindamycin, tabcap, 300 mg |
| Second trimester - uncomplicated | Paracetamol 500mg<br>Lumefantrine 120mg/Artemether 20mg |
| **Complicated (severe)** | |
| Adults | Injectable artesunate |
| Children | Injectable artesunate |
| Pregnant women | Quinine dihydrochloride 300mg/ml<br>Quinine sulphate 300mg<br>Clindamycin, tabcap, 300 mg<br>Dextrose (glucose) 5% |

Table 4: Treatment options for malaria following the Malawi Standard Treatment Guidelines 2015 ([40])

| Disease stage | Description | DALY weights |
|---|---|---|
| HIV infection | Symptomatic HIV without anaemia, HIV cases, symptomatic, pre-AIDS, "has weight loss, fatigue, and frequent infections. | 0.274 (0.184-0.377) |
| AIDS | AIDS without antiretroviral treatment without anaemia, AIDS cases, not receiving ARV treatment, has severe weight loss, weakness, fatigue, cough and fever, and frequent infections, skin rashes and diarrhoea | 0.582 (0.406-0.743) |
| Latent TB | Latent tuberculosis infection, Asymptomatic | – |
| Active TB, HIV-negative | Drug-susceptible or multidrug-resistant tuberculosis, not HIV infected, has a persistent cough and fever, is short of breath, feels weak, and has lost a lot of weight | 0.333 (0.224-0.454) |
| Active TB, HIV-positive | Drug-susceptible or multidrug-resistant HIV/AIDS - Tuberculosis without anaemia, HIV infected, has a persistent cough and fever, shortness of breath, night sweats, weakness and fatigue and severe weight loss | 0.408 (0.274-0.549) |
| Asymptomatic malaria parasitaemia | Asymptomatic | – |
| Moderate malaria with mild anaemia | Infectious disease, acute episode, moderate, with mild anaemia. combined DALY weight: i) has a fever and aches, and feels weak, which causes some difficulty with daily activities ii) feels slightly tired and weak at times, but this does not interfere with normal daily activities. | 0.054 (0.034-0.079) |
| Severe malaria | Infectious disease, acute episode. Has a high fever and pain, and feels very weak, which causes great difficulty with daily activities. | 0.133 (0.088-0.19) |

Table 5: DALY weights associated with HIV, TB and malaria infections. DALY weights for drug-susceptible and multidrug-resistant TB strains are assumed to be identical.[13]

| Interaction | Value | Source |
|---|---|---|
| **Interactions with HIV** | | |
| RR HIV with Schistosoma haematobium | 2.3 | [41] |
| RR TB with untreated HIV | 5 | Assumption |
| RR TB with AIDS | 26 | [31] |
| RR TB relapse with untreated HIV | 4.7 | [42] |
| Proportion of TB cases smear-positive with untreated HIV | 0.35 | [5] |
| RR clinical malaria in children under 5 years with untreated HIV | 1.7 | [35] |
| RR clinical malaria in people aged over 5 years with untreated HIV | 2.6 | [35] |
| RR clinical malaria with pregnant women with untreated HIV | 3.96 | [37] |
| RR severe malaria in children under 5 years with untreated HIV | 9.69 | [43] |
| RR severe malaria in people aged over 5 years with untreated HIV | 2.68 | [43] |
| RR severe malaria with pregnant women with untreated HIV | 2.8 | [38] |
| RR acquiring ALRI for children who are HIV positive | 4.15 | [44] |
| RR ALRI death with untreated HIV | 1.5 | Assumption |
| RR moderate to severe diarrhoea in children with untreated HIV | 5.6 | [45] |
| RR diarrhoea death in children with untreated HIV | 5.05 | [45] |
| RR anaemia in pregnant women with untreated HIV | 4.19 | [46] |
| RR stunting in children with untreated HIV | 1.5 | Assumption |
| RR progression to severe stunting in children with untreated HIV | 1.3 | Assumption |
| RR depression with HIV | 1.99 | [47] |
| RR cardiovascular / cerebrovascular events with untreated HIV | 1.61 | [48] |
| RR non-AIDS cancers with untreated HIV | 2.0 | [49] |
| **Interactions with TB** | | |
| RR TB with diabetes | 1.5 | [50] |
| RR TB relapse with diabetes | 1.86 | [51] |
| RR TB death with diabetes | 1.51 | [51] |
| RR TB with untreated HIV and diabetes | 1.5 | [50] |
| **Interactions with malaria** | | |
| RR anaemia in pregnant women with malaria | 1.45 | [52] |
| RR preterm labour with malaria | 3.08 | [53] |
| RR stillbirth with malaria | 1.81 | [54] |

Table 6: Key interactions between HIV, TB, malaria, and other disorders included in the TLO model. RR = relative risk.

| Appt type | Clinical | Nursing | Pharmacy | Radiography | Notes |
|---|---|---|---|---|---|
| HIV Prevention Circumcision | 18.75 | 6 | | | |
| TB Test Screening | 27 | 18 | 9.45 | | Assume footprint of Over5OPD |
| HIV Prevention Infant | 2.5 | 28.75 | 3 | | Includes VCTNeg + Peds |
| HIV Test | | 20.75 | | | Weighted mean of 20mins VCTNeg and 35 mins VCTPos assuming constant 5% testing yield |
| HIV Treatment | 2.5 | 8.75 | 3 | | All EstNonCom |
| HIV Palliative Care | 263 | 651 | 103 | | 1*IPAdmission, 17*InpatientDays, level2 |
| TB Test X-ray | | | | 18 | District hospital |
| TB Treatment | 15.25 | 8.75 | 5 | | |
| Malaria Treatment | 27 | 18 | 9.45 | | Assume footprint of Over5OPD |
| Malaria Test | 27 | 18 | 9.45 | | Assume footprint of Over5OPD |
| Malaria Treatment Complicated | 149 | 216 | 48.7 | | Includes Over5OPD at district hospital, 1 inpatient admission and 5 inpatient days |
| TB Test FollowUp | 11 | 3 | 4 | | |
| TB Palliative Care | 139.5 | 309 | 50.75 | | Includes 1 inpatient admission and 7.5 inpatient days at referral hospital |
| Malaria Prevention IPTp | 27 | 18 | 9.45 | | Assume footprint of Over5OPD |
| TB Prevention IPT | 27 | 18 | 9.45 | | Assume footprint of Over5OPD |
| HIV Prevention PrEP | | 20 | 5 | | Includes a pharmacy appointment and VCTNeg |

Table 7: The patient-facing time required for specific healthcare services in minutes. The patient-facing time is an example of the time taken for a standalone appointment to deliver each service. In practice, and in the simulation, these services may be integrated with other services, reducing the human resources requirement. Abbreviations: Over5OPD - patient over 5 years outpatient appointment; VCTNeg - voluntary testing and counselling for HIV-negative person; Peds - paediatric appointment; VCTPos - voluntary testing and counselling for HIV-positive person; EstNonCom - established non-medically-complex HIV patient; IPAdmission - inpatient admission.

| | Death Rate in Interval | | | |
|---|---|---|---|---|
| Age range | Actual | | No HTM services | |
| | Males | Females | Males | Females |
| 0 | 0.0402 | 0.0465 | 0.0813 | 0.0850 |
| 1-4 | 0.0068 | 0.0060 | 0.0273 | 0.0254 |
| 5 − 9 | 0.0019 | 0.0020 | 0.0022 | 0.0017 |
| 10 − 14 | 0.0012 | 0.0008 | 0.0024 | 0.0011 |
| 15 − 19 | 0.0010 | 0.0010 | 0.0050 | 0.0036 |
| 20 − 24 | 0.0038 | 0.0021 | 0.0081 | 0.0076 |
| 25 − 29 | 0.0028 | 0.0032 | 0.0080 | 0.0070 |
| 30 − 34 | 0.0054 | 0.0055 | 0.0098 | 0.0102 |
| 35 − 39 | 0.0063 | 0.0041 | 0.0144 | 0.0190 |
| 40 − 44 | 0.0071 | 0.0027 | 0.0206 | 0.0235 |
| 45 − 49 | 0.0102 | 0.0059 | 0.0176 | 0.0261 |
| 50 − 54 | 0.0132 | 0.0088 | 0.0330 | 0.0286 |
| 55 − 59 | 0.0179 | 0.0094 | 0.0221 | 0.0186 |
| 60 − 64 | 0.0357 | 0.0187 | 0.0285 | 0.0250 |
| 65 − 69 | 0.0428 | 0.0189 | 0.0401 | 0.0360 |
| 70 − 74 | 0.0863 | 0.0493 | 0.0649 | 0.0358 |
| 75 − 79 | 0.0513 | 0.0611 | 0.0715 | 0.0673 |
| 80 − 84 | 0.0679 | 0.0598 | 0.1547 | 0.0700 |
| 85 − 89 | 0.2262 | 0.1376 | 0.1175 | 0.1621 |
| 90 + | 0.2230 | 0.1211 | 0.1933 | 0.1813 |

Table 8: The annual age-specific mortality rates estimated using the simulated numbers of deaths per population in 2019. Outputs are shown from one run.

| Cause of death | Actual scenario | No HIV services | No TB services | No malaria services | No HTM services |
|---|---|---|---|---|---|
| AIDS inc TB | 210.9 (207.9 - 222.2) | 779.9 (770.8 - 785.1) | 238.1 (228.7 - 243.2) | 210.3 (207.8 - 218.1) | 790.4 (786.5 - 801.3) |
| TB excl HIV | 58.7 (57.7 - 63.1) | 63.6 (63.5 - 64.9) | 157.3 (152.0 - 164.0) | 59.0 (56.7 - 66.9) | 154.0 (151.9 - 159.5) |
| Malaria | 120.9 (113.1 - 121.8) | 116.5 (111.7 - 118.8) | 116.3 (114.7 - 125.5) | 538.8 (526.1 - 546.1) | 537.4 (527.1 - 542.6) |
| All causes | 1,327.7 (1,313.9 - 1,346.0) | 1,899.4 (1,881.6 - 1,915.3) | 1,440.9 (1,429.5 - 1,448.5) | 1,733.8 (1,726.2 - 1,745.6) | 2,412.8 (2,388.1 - 2,434.6) |
| Life expectancy males, years | 61.7 (61.1 - 62.3) | 53.4 (52.3 - 54.2) | 60.6 (60.1 - 61.0) | 57.3 (56.8 - 57.8) | 48.2 (47.6 - 49.0) |
| Life expectancy females, years | 66.1 (65.6 - 66.8) | 54.9 (54.0 - 55.2) | 65.5 (64.8 - 66.9) | 61.7 (60.6 - 62.9) | 50.4 (49.7 - 51.2) |

Table 9: Total deaths (thousands) estimated under each scenario and estimated life expectancy from birth for males and females using the all-cause mortality rates in 2019.

| | **Actual scenario** | **No HIV services** | **No TB services** | **No malaria services** | **No HTM services** |
|---|---|---|---|---|---|
| AIDS | 1.291 (1.274-1.364) | 4.853 (4.788-4.888) | 1.464 (1.408-1.498) | 1.301 (1.287-1.352) | 4.992 (4.961-5.06) |
| COPD | 0.149 (0.138-0.157) | 0.139 (0.127-0.152) | 0.132 (0.12-0.14) | 0.143 (0.127-0.15) | 0.138 (0.124-0.147) |
| Cancer (Bladder) | 0.055 (0.042-0.06) | 0.052 (0.043-0.055) | 0.046 (0.037-0.06) | 0.047 (0.039-0.054) | 0.045 (0.043-0.049) |
| Cancer (Breast) | 0.075 (0.057-0.078) | 0.074 (0.061-0.078) | 0.071 (0.063-0.08) | 0.069 (0.065-0.089) | 0.073 (0.056-0.079) |
| Cancer (Oesophagus) | 0.05 (0.038-0.054) | 0.044 (0.042-0.06) | 0.046 (0.037-0.054) | 0.049 (0.038-0.054) | 0.046 (0.037-0.059) |
| Cancer (Other) | 0.436 (0.43-0.441) | 0.464 (0.457-0.488) | 0.423 (0.42-0.46) | 0.435 (0.426-0.447) | 0.48 (0.457-0.492) |
| Cancer (Prostate) | 0.047 (0.045-0.05) | 0.052 (0.044-0.055) | 0.05 (0.041-0.062) | 0.057 (0.05-0.059) | 0.057 (0.05-0.062) |
| Childhood Diarrhoea | 0.195 (0.177-0.206) | 0.252 (0.246-0.263) | 0.177 (0.172-0.185) | 0.179 (0.168-0.185) | 0.245 (0.229-0.249) |
| Congenital birth defects | 0.027 (0.018-0.029) | 0.025 (0.022-0.031) | 0.025 (0.019-0.034) | 0.026 (0.019-0.03) | 0.022 (0.02-0.025) |
| Depression / Self-harm | 0.068 (0.066-0.077) | 0.073 (0.062-0.081) | 0.073 (0.065-0.087) | 0.073 (0.065-0.077) | 0.069 (0.066-0.081) |
| Diabetes | 0.1 (0.096-0.107) | 0.106 (0.093-0.11) | 0.103 (0.092-0.108) | 0.102 (0.1-0.121) | 0.096 (0.083-0.108) |
| Epilepsy | 0.014 (0.01-0.017) | 0.015 (0.009-0.02) | 0.012 (0.01-0.02) | 0.012 (0.009-0.017) | 0.016 (0.012-0.017) |
| Heart Disease | 0.39 (0.371-0.419) | 0.383 (0.35-0.392) | 0.368 (0.328-0.396) | 0.384 (0.36-0.408) | 0.371 (0.36-0.391) |
| Kidney Disease | 0.069 (0.062-0.082) | 0.064 (0.061-0.067) | 0.069 (0.064-0.075) | 0.068 (0.06-0.077) | 0.067 (0.062-0.079) |
| Lower respiratory infections | 0.802 (0.794-0.812) | 0.853 (0.837-0.874) | 0.806 (0.799-0.836) | 0.796 (0.776-0.812) | 0.853 (0.824-0.889) |
| Malaria | 0.74 (0.694-0.747) | 0.726 (0.694-0.74) | 0.716 (0.706-0.772) | 3.336 (3.259-3.383) | 3.392 (3.328-3.423) |
| Maternal Disorders | 0.131 (0.121-0.15) | 0.133 (0.128-0.146) | 0.138 (0.123-0.145) | 0.132 (0.122-0.141) | 0.137 (0.132-0.162) |
| Measles | 0.095 (0.083-0.098) | 0.089 (0.087-0.099) | 0.094 (0.081-0.098) | 0.086 (0.078-0.095) | 0.089 (0.081-0.103) |
| Neonatal Disorders | 0.749 (0.729-0.763) | 0.743 (0.724-0.762) | 0.726 (0.722-0.757) | 0.786 (0.748-0.791) | 0.787 (0.738-0.829) |
| Stroke | 0.321 (0.307-0.33) | 0.313 (0.299-0.337) | 0.32 (0.311-0.348) | 0.324 (0.299-0.341) | 0.304 (0.286-0.335) |
| TB (non-AIDS) | 0.359 (0.354-0.387) | 0.396 (0.395-0.403) | 0.967 (0.936-1.009) | 0.366 (0.351-0.414) | 0.972 (0.959-1.007) |
| Transport Injuries | 0.285 (0.272-0.316) | 0.293 (0.281-0.307) | 0.3 (0.279-0.311) | 0.287 (0.276-0.321) | 0.28 (0.268-0.31) |

Table 10: Median mortality rates per 1000 person-years with 95% uncertainty intervals by scenario. Mortality rates are calculated as the numbers of cause-specific deaths divided by person-years for each run and then aggregated by scenario.

| | **Single programme estimate** | **Joint programme estimate** |
|---|---|---|
| HIV/AIDS | 562,700 (555,900 - 574,300) | 579,300 (570,900 586,100) |
| TB | 95,400 (91,700 - 105,700) | 94,200 (90,400 100,900) |
| Malaria | 420,300 (408,400 - 424,900) | 416,100 (414,000 420,800) |
| Total | 1,078,400 | 1,089,600 |

Table 11: Total deaths averted due by HIV/AIDS, TB and malaria cause rounded to the nearest 100. Single programme estimates are shown for the hypothetical single programme exclusion scenarios (e.g. HIV/AIDS deaths estimated had no HIV services been available, TB deaths had no TB services been available) and then calculated jointly had no HTM services been available.

| | Median % change | Lower 95% bound | Upper 95% bound |
|---|---|---|---|
| AIDS | 234.273 | 224.285 | 242.023 |
| COPD | -4.213 | -16.683 | 0.340 |
| Cancer (Bladder) | -12.662 | -22.606 | 0.744 |
| Cancer (Breast) | -1.037 | -11.928 | 6.500 |
| Cancer (Oesophagus) | 4.688 | -9.089 | 17.675 |
| Cancer (Other) | 12.070 | 6.269 | 14.409 |
| Cancer (Prostate) | 19.403 | 1.631 | 24.451 |
| Childhood Diarrhoea | 24.946 | 19.242 | 37.806 |
| Congenital birth defects | -8.606 | -17.801 | 15.753 |
| Depression / Self-harm | -3.730 | -4.685 | -1.883 |
| Diabetes | -2.264 | -10.395 | 3.422 |
| Epilepsy | 9.114 | -4.739 | 13.732 |
| Heart Disease | -6.126 | -9.644 | 0.017 |
| Kidney Disease | 7.553 | 1.378 | 18.499 |
| Lower back pain | -2.068 | -2.836 | -1.740 |
| Lower respiratory infections | 6.790 | 2.730 | 9.678 |
| Malaria | 392.041 | 383.975 | 409.748 |
| Maternal Disorders | 5.829 | -9.952 | 31.004 |
| Measles | 8.674 | 0.507 | 15.648 |
| Neonatal Disorders | 4.993 | -1.398 | 11.174 |
| Stroke | -0.802 | -2.384 | 2.987 |
| Schistosomiasis | -2.263 | -11.432 | 7.383 |
| TB (non-AIDS) | 174.110 | 149.131 | 183.231 |
| Transport Injuries | -2.569 | -4.332 | 6.582 |

Table 12: Percentage change in DALYs incurred per person-year (median, lower and upper 95% uncertainty intervals) estimated when no HTM services are available compared with the Actual scenario.

| DALYS per PY | Actual | No HIV services | No TB services | No malaria services | No HTM services |
|---|---|---|---|---|---|
| AIDS inc TB | 0.088<br>0.087 - 0.093 | 0.291<br>0.287 - 0.294 | 0.097<br>0.093 - 0.099 | 0.089<br>0.088 - 0.092 | 0.297<br>0.294 - 0.301 |
| TB excl HIV | 0.021<br>0.020 - 0.022 | 0.023<br>0.023 - 0.024 | 0.055<br>0.054 - 0.058 | 0.021<br>0.021 - 0.024 | 0.056<br>0.056 - 0.058 |
| Malaria | 0.060<br>0.056 - 0.061 | 0.058<br>0.056 - 0.059 | 0.058<br>0.057 - 0.062 | 0.288<br>0.282 - 0.291 | 0.292<br>0.288 - 0.296 |
| Other causes | 0.371<br>0.368 - 0.379 | 0.383<br>0.378 - 0.386 | 0.370<br>0.369 - 0.377 | 0.373<br>0.370 - 0.377 | 0.383<br>0.377 - 0.394 |

Table 13: Total DALYs incurred grouped by AIDS, TB (non-AIDS), malaria and other causes (non-HTM) during the simulation divided by person-years. Values are summarised across the runs of each draw.

| | Actual scenario | No HIV services | No TB services | No malaria services | No HTM services |
|---|---|---|---|---|---|
| AIDS | 14559.4 (14242.3-15094.7) | 46686.4 (46203.5-47171.3) | 15644.8 (15128.9-16126.3) | 14428.4 (14173.6-14868.9) | 47103.1 (46611.1-47624.8) |
| COPD | 894.6 (828.9-937.1) | 830.1 (782.2-864.8) | 816.5 (774.1-844.9) | 849.0 (810.5-889.4) | 804.5 (757.9-847.0) |
| Cancer (Bladder) | 425.2 (353.9-491.1) | 407.8 (349.5-446.9) | 397.5 (310.8-476.8) | 389.6 (330.4-433.5) | 359.6 (324.8-399.7) |
| Cancer (Breast) | 518.4 (402.2-564.8) | 508.2 (424.9-578.5) | 528.3 (455.6-599.0) | 522.8 (440.4-661.7) | 485.9 (382.2-565.5) |
| Cancer (Oesophagus) | 297.4 (249.5-343.5) | 307.6 (259.8-375.6) | 294.7 (247.0-350.3) | 296.6 (246.4-333.8) | 297.3 (237.5-365.1) |
| Cancer (Other) | 2565.8 (2531.2-2613.0) | 2790.2 (2712.2-2888.1) | 2554.5 (2467.3-2731.7) | 2555.4 (2451.6-2637.4) | 2774.8 (2629.5-2861.8) |
| Cancer (Prostate) | 271.4 (259.1-297.0) | 288.4 (259.0-319.2) | 285.5 (246.8-325.1) | 305.3 (274.4-326.6) | 306.1 (265.6-345.6) |
| Childhood Diarrhoea | 2774.8 (2575.9-2980.3) | 3635.8 (3516.5-3750.7) | 2568.6 (2477.6-2678.1) | 2549.3 (2413.1-2654.1) | 3411.6 (3222.6-3506.4) |
| Congenital birth defects | 360.4 (255.5-429.3) | 376.7 (313.7-446.8) | 373.2 (273.8-491.9) | 350.7 (275.7-430.0) | 321.8 (286.1-351.5) |
| Depression / Self-harm | 5834.2 (5779.9-5885.0) | 5533.7 (5475.4-5577.2) | 5829.9 (5734.2-5940.0) | 5871.3 (5818.7-5923.2) | 5468.9 (5405.9-5563.0) |
| Diabetes | 517.7 (502.2-535.4) | 529.1 (492.6-562.4) | 524.6 (489.9-568.5) | 551.4 (499.5-627.0) | 482.6 (446.6-513.0) |
| Epilepsy | 570.1 (537.7-603.9) | 582.2 (525.3-635.1) | 576.2 (532.0-655.7) | 561.2 (515.2-606.4) | 581.4 (541.3-599.6) |
| Heart Disease | 1327.4 (1249.5-1407.4) | 1244.0 (1175.0-1314.0) | 1202.9 (1027.9-1323.5) | 1278.2 (1191.2-1338.3) | 1225.6 (1209.5-1248.6) |
| Kidney Disease | 221.0 (202.2-263.4) | 205.4 (193.0-216.5) | 229.2 (210.5-255.3) | 232.0 (208.5-260.8) | 235.7 (213.3-260.3) |
| Lower back pain | 546.4 (539.6-551.0) | 525.0 (518.2-530.0) | 542.5 (538.4-544.7) | 547.5 (538.5-551.9) | 518.9 (512.3-524.1) |
| Lower respiratory infections | 11645.1 (11495.0-11774.4) | 12166.3 (11964.6-12483.4) | 11782.4 (11525.0-12082.3) | 11416.6 (11149.4-11658.6) | 12050.3 (11607.1-12519.4) |
| Malaria | 9652.1 (9194.5-9967.9) | 9271.4 (9015.0-9491.3) | 9611.7 (9264.9-10151.3) | 46349.2 (45500.5-46997.2) | 46314.2 (45556.6-46903.1) |
| Maternal Disorders | 1388.3 (1275.8-1563.6) | 1388.4 (1313.4-1502.3) | 1422.2 (1284.7-1519.0) | 1365.4 (1257.1-1421.4) | 1466.6 (1348.9-1634.5) |
| Measles | 1199.5 (1089.9-1311.4) | 1211.6 (1138.6-1293.7) | 1245.8 (1100.1-1311.8) | 1150.6 (1055.2-1260.7) | 1261.7 (1185.7-1376.5) |
| Neonatal Disorders | 11053.2 (10815.4-11300.3) | 10857.3 (10539.8-11104.8) | 10833.6 (10661.1-11175.8) | 11346.7 (10982.8-11586.9) | 11336.6 (10612.7-11926.9) |
| Other | 14014.8 (13657.9-14535.1) | 13742.3 (13450.9-14172.7) | 14114.5 (13897.0-14302.1) | 13785.2 (13260.7-14045.8) | 13529.6 (12906.8-14094.4) |
| Stroke | 93.0 (90.7-94.3) | 92.4 (90.5-93.3) | 92.2 (89.8-94.6) | 90.3 (87.4-93.4) | 90.4 (89.1-93.0) |
| Schistosomiasis | 1263.6 (1207.7-1298.7) | 1240.8 (1192.5-1328.7) | 1289.1 (1210.7-1418.3) | 1274.5 (1208.1-1340.9) | 1197.0 (1111.9-1280.0) |
| TB (non-AIDS) | 3478.9 (3343.2-3635.6) | 3771.0 (3710.7-3818.6) | 9036.4 (8700.6-9427.4) | 3549.9 (3366.2-3898.6) | 9018.9 (8813.7-9250.5) |
| Transport Injuries | 2999.1 (2838.8-3239.5) | 3033.0 (2872.3-3155.5) | 3049.9 (2877.3-3248.8) | 3006.9 (2822.6-3325.7) | 2896.2 (2803.4-3035.7) |

Table 14: Median DALYs incurred (thousands) with 95% uncertainty intervals by scenario.

| | Actual scenario | | | No HTM | | |
|---|---|---|---|---|---|---|
| | Median | Lower | Upper | Median | Lower | Upper |
| ALRI | 6,629.2 | 6,526 | 6,745 | 6,817.3 | 6,755.4 | 6,959.8 |
| Antenatal Care | 24,532.6 | 24,291.4 | 24,769.4 | 23,770.8 | 23,618.7 | 24,007.6 |
| Bladder Cancer | 202.7 | 195.3 | 222.4 | 179.1 | 161.1 | 182.3 |
| Breast Cancer | 204.1 | 171.9 | 226 | 207.3 | 157 | 223.4 |
| CardioMetabolic Disorders | 10,880.6 | 10,677.1 | 11,124.9 | 11,106.6 | 10,965.3 | 11,181.4 |
| Contraception | 345,124.1 | 337,539.4 | 352,157.7 | 331,995 | 329,842.8 | 339,809.6 |
| COPD | 316.7 | 293.5 | 332.3 | 299.6 | 286.5 | 308.3 |
| Delivery Care | 5,057.3 | 5,005.7 | 5,121.3 | 4,908.3 | 4,874.1 | 4,937.7 |
| Depression | 325 | 297 | 346.1 | 323.9 | 312.8 | 327.2 |
| Diarrhoea | 17,076.4 | 16,938.2 | 17,129.8 | 34,356.3 | 34,188.8 | 34,531.3 |
| EPI | 99,368 | 98,596.2 | 100,377.8 | 96,390 | 95,771.8 | 97,010.7 |
| Epilepsy | 25,156.9 | 23,565.4 | 25,599.6 | 25,615 | 24,978 | 25,931.2 |
| HIV | 42,917 | 42,805 | 43,186 | 799.2 | 793 | 807.6 |
| Malaria | 98,177.9 | 97,545.6 | 98,498.8 | 541.3 | 531 | 547 |
| Measles | 2,617.8 | 2,610.5 | 2,642.7 | 2,529.1 | 2,512.1 | 2,555.9 |
| Oesophageal Cancer | 174.2 | 155.8 | 178.6 | 166.4 | 143.7 | 197.7 |
| Other Adult Cancer | 436.4 | 419.5 | 500.9 | 447.5 | 434.4 | 473.1 |
| Postnatal Care | 9,958.4 | 9,876 | 10,135.5 | 9,725.9 | 9,700.1 | 9,840.9 |
| Prostate Cancer | 120.5 | 109.9 | 129.5 | 119.2 | 104.3 | 125.3 |
| RTI | 309,101.4 | 294,622.4 | 319,903.4 | 299,356.8 | 295,570.2 | 308,390.6 |
| Schistosomiasis | 712.3 | 699.8 | 715.3 | 1,137.3 | 1,114.2 | 1,160 |
| TB | 62,320.4 | 62,278.3 | 62,525.9 | 155.2 | 153.2 | 161.2 |
| Undernutrition | 325.6 | 315.9 | 332.7 | 629.6 | 616.6 | 662.8 |
| First Attendance Emergency | 8,699.2 | 8,447.5 | 8,802.8 | 9,086.5 | 8,983.4 | 9,175.3 |
| First Attendance NonEmergency | 33,522.6 | 33,417.7 | 33,553.2 | 53,997.7 | 53,792.5 | 54,103.8 |

Table 15: The numbers of appointments (in thousands) classified by disease programme for the Actual scenario and if no HTM services had been available. The remaining HIV, TB and malaria services in the "No HTM" results relate to end-of-life or palliative care. First Attendance appointments refer to general outpatient appointments which are stratified into emergency and non-emergency appointments. The median values of 5 runs are shown for each scenario along with the lower (2.5th percentile) and upper (97.5th percentile) bounds.

| | Median | Service delivery |
|---|---|---|
| Testing services | 157.029<br>(156.364 - 157.545) | Delivered through community, outreach or outpatient clinics.<br>Often integrated with other services. |
| Treatment and follow-up services | 23.184<br>(23.092 - 23.288) | All are assumed to occur at outpatient clinics<br>at the primary care level. |
| Preventive services | 22.659<br>(22.468 - 22.922) | Delivered through community, outreach or outpatient clinics.<br>Often integrated with other services. |
| Inpatient admissions | 0.559<br>(0.548 - 0.561) | Inpatient care offered through district, regional or central hospitals.<br>Each count refers to one patient admission. |

Table 16: Estimated numbers of health system services required in millions for the provision of HTM care.

| Cause of death | Actual scenario | Include HIV prog only | Include TB prog only | Include malaria prog only | No HTM services |
|---|---|---|---|---|---|
| AIDS inc TB | 210.9 (207.9 - 222.2) | 322.2 (315.1 - 330.2) | 816.6 (812.9 - 829.4) | 847.0 (832.3 - 847.5) | 790.4 (786.5 - 801.3) |
| TB excl HIV | 58.7 (57.7 - 63.1) | 156.1 (147.1 - 161.2) | 59.3 (57.2 - 66.4) | 151.5 (150.4 - 157.2) | 154.0 (151.9 - 159.5) |
| Malaria | 120.9 (113.1 - 121.8) | 541.4 (523.6 - 545.8) | 523.3 (519.9 - 536.0) | 114.8 (110.2 - 117.4) | 537.4 (527.1 - 542.6) |
| All causes | 1,327.7 (1,313.9 - 1,346.0) | 1,947.8 (1,9045.0 - 1,56.9) | 2,334.9 (2,324.9 - 2,349.2) | 2,040.7 (2,029.6 - 2,043.8) | 2,412.8 (2,388.1 - 2,434.6) |
| AIDS mortality rates per 1000py | 1.29 (1.27 - 1.36) | 2.01 (1.97 - 2.06) | 5.15 (5.12 - 5.24) | 5.30 (5.22 - 5.32) | 4.99 (4.96 - 5.06) |
| TB mortality rates per 1000py | 0.36 (0.35 - 0.39) | 0.98 (0.92 - 1.01) | 0.37 (0.36 - 0.42) | 0.95 (0.94 - 0.99) | 0.97 (0.96 - 1.01) |
| Malaria mortality rates per 1000py | 0.74 (0.69 - 0.75) | 3.38 (3.27 - 3.41) | 3.30 (3.29 - 3.38) | 0.72 (0.69 - 0.74) | 3.39 (3.33 - 3.43) |
| HTM mortality rates per 1000py | 2.41 (2.38 - 2.43) | | 3.13 (3.05 - 3.14) | | 9.38 (9.30 - 9.41) |
| Life expectancy males, years | 61.7 (61.1 - 62.3) | 55.5 (54.8 - 56.6) | 50.0 (49.7 - 50.4) | 53.0 (52.7 - 53.2) | 48.2 (47.6 - 49.0) |
| Life expectancy females, years | 66.1 (65.6 - 66.8) | 59.7 (58.2 - 60.9) | 51.5 (50.8 - 52.5) | 55.4 (54.4 - 56.0) | 50.4 (49.7 - 51.2) |

Table 17: Numbers of deaths (thousands) due to HIV/AIDS, TB, malaria and all causes; mortality rates due to HIV/AIDS, TB, malaria and HTM combined per 1000 person-years; life expectancy from birth estimated under the additional scenarios. Life expectancy for males and females is calculated using the all-cause mortality rates in 2019.

| Cause of death | Include HIV prog only | Include TB prog only | Include malaria prog only | No HTM services |
|---|---|---|---|---|
| AIDS inc TB | 471.5 (460.1 - 477.4) | -27.4 (-37.5 - -21.1) | -56.6 (-60.2 - -37.1) | 579.3 (570.9 - 586.1) |
| TB excl HIV | -4.25 (-8.03 - 12.0) | 94.7 (87.8 - 100.9) | 3.66 ( -5.32 - 8.00) | 94.2 (90.4 - 100.9) |
| Malaria | -5.04 (-14.9 - 19.0) | 6.73 (2.61 - 18.6) | 420.5 (416.7 - 427.6) | 416.1 (414.0 - 420.8) |
| HTM | | 984.9 (973.7 - 999.7) | | 1093.7 (1077.5 - 1100.3) |

Table 18: Numbers of deaths averted (thousands) due to the HIV/AIDS, TB, malaria programmes separately and when no HTM services are available (main analysis). The estimate of HTM deaths averted using the single programme estimates is the sum of the HIV/AIDS, TB and malaria deaths averted through the single programmes (highlighted in blue).
